# Transferability and interpretability of the sepsis prediction models in the intensive care unit

**DOI:** 10.1101/2021.05.13.21256281

**Authors:** Qiyu Chen, Ranran Li, ChihChe Lin, Chiming Lai, Dechang Chen, Hongping Qu, Yaling Huang, Wenlian Lu, Yaoqing Tang, Lei Li

## Abstract

**Objective:** We aimed to develop an early warning system for real-time sepsis prediction in the ICU by equipping with interpretation analysis and transfer learning tools to improve the feasibility to deploy the sepsis prediction system, particularly to target cohorts.

**Design:** Retrospective and observational study.

**Setting:** Medical Information Mart for Intensive Care (MIMIC) dataset, the private Historical Database of local Ruijin Hospital (HDRJH), and data collected from Ruijin real-world study.

**Patients:** 6891 patients from MIMIC dataset and 453 patients from HDRJH for model development and 67 cases from Ruijin real-world data for model evaluation.

**Interventions:** None.

**Measurements and Main Results:** Light Gradient Boosting Machine (LightGBM) and multilayer perceptron (MLP) were trained on MIMIC dataset and then finetuned on HDRJH using transfer learning technique. Ultimately, the performance of the sepsis prediction system was further evaluated in the real-world study in the ICU of the target Ruijin Hospital. The area under the receiver operating characteristic curves (AUCs) for LightGBM and MLP models derived from MIMIC were 0.98–0.98 and 0.95–0.96 respectively on MIMIC dataset, and, in comparison, 0.82–0.86 and 0.84–0.87 respectively on HDRJH, from 1–5h preceding. After transfer learning and ensemble learning, the AUCs of the final ensemble model were enhanced to 0.94–0.94 on HDRJH and to 0.86–0.9 in the real-world study in the ICU of the target Ruijin Hospital. In addition, the Shapley additive explanation (SHAP) analysis illustrated the importance of age, antibiotics, net balance, and ventilation for sepsis prediction, making the model interpretable.

**Conclusions:** Our machine learning model allows accurate real-time prediction of sepsis within 5-h preceding. Transfer learning can effectively improve the feasibility to deploy the prediction model in the target cohort, effectively ameliorating the model performance for external validation. SHAP analysis may illuminate the importance of optimizing antibiotic use and restricting fluid management.

**Trial registration:** NCT05088850 (retrospectively registered).

**Key Points:** *Question:* We aimed to develop an early warning system for real-time sepsis prediction in the ICU and to improve the feasibility to deploy the system to target cohorts.

*Findings:* Transfer learning technique effectively enhanced the AUCs for LightGBM and MLP models on the target cohort, HDRJH, from 0.82–0.86 and 0.84–0.87 to 0.93-0.94 and 0.92-0.93 for 1-5 hour preceding. Additionally, SHAP analysis illuminated the importance of optimizing antibiotic use and restricting fluid management.

*Meaning:* Transfer learning can improve the feasibility to deploy the prediction model to the target cohort, and SHAP analysis made the prediction model interpretable.

---

Sepsis, an infection-induced syndrome of physiological, pathological, and biochemical abnormalities, is a global healthcare issue associated with unacceptably high mortality and long-term morbidity among patients in the intensive care unit (ICU) (1,2), and is responsible for a substantial cost burden on health care resources (3). Early detection and timely administration of appropriate antibiotics are important for improving the prognosis and survival of septic patients (4). However, nonspecific symptoms of sepsis may cause delayed diagnosis and intervention, leading to the high mortality of septic patients (5).

Machine learning has emerged as a promising tool for the early detection of sepsis occurrence based on electronic medical records, laboratory data, and biomedical signals (6-14). Several prospective studies have shown that implementation of machine learning-based sepsis prediction algorithms can reduce in-hospital mortality and length of stay (15,16). Except for the excellent prediction performance, the translation of these risk prediction models into clinical practice requires external independent validation to determine the generalizability of the model to different cohorts (17). However, most of the newly proposed risk prediction models have discriminatory performance when applied to external samples (18). Re-training of the prediction model on local datasets in the target medical center might enhance the predictive accuracy in the specific situation (19). Transfer learning as a re-training approach has been reported to improve model performance when the dataset is small in the target medical center (20,21). Additionally, the interpretability of machine learning models reflects the extent to which the decision-making process of the model can be understood and accepted in clinical practice. The lack of interpretability for most available prediction models is currently the major barrier to their clinical adoption (22,23). The objective of this study is to develop an interpretable early warning system (named SEPRES, Sepsis PREdiction System) for real-time sepsis prediction in the ICU and to improve its generalizability to the target medical center through transfer learning technique.

## Methods

SEPRES includes a data integration system equipped with a sepsis early warning module. The data integration system collects, stores, processes, and displays medical data. The sepsis early warning module included a sepsis prediction model and an interpretative tool. The sepsis prediction model is an ensemble of multiple machine learning models. The interpretative tool provides information on how the model works by assigning importance to the input features. Our study complies with the relevant reporting guidelines, namely the Transparent Reporting of a multivariable prediction model for Individual Prognosis Or Diagnosis (TRIPOD) statement (24). The study protocol complies with the Declaration of Helsinki, as revised in 2013, and was approved by the Ruijin Hospital Ethics Committee (No. 2020 [140], approved on October 27, 2020, study title: The early. Prediction of sepsis occurrence in ICU). Written informed consent was obtained from individual or guardian participants.

### Data acquisition

#### Data sources

Our study used the Medical Information Mart for Intensive Care (MIMIC-III) database (version 1.4) (25) and the private Historical Database of Ruijin Hospital (HDRJH). MIMIC encompasses 61532 patients admitted to the ICU at Beth Israel Deaconess Medical Center in Boston from 2001 to 2012, and HDRJH encompasses approximately 1777 patients from 2011 to 2019. In addition to retrospective data, we also collected predictions of consecutive 67 patients from the SEPRES system running in the ICU at Ruijin (RJ) Hospital between February 2021 and June 2021 as a validation of the model in the real world.

#### Sepsis definitions

We defined sepsis according to the definition of the Third International Consensus for sepsis (Sepsis-3) (2), combining suspected infection and SOFA score.

#### Feature extraction

We extracted 78 and 63 patient variables from the MIMIC and HDRJH, respectively. After data cleaning, we extracted these variables as features, i.e., maximum, average, median, and minimum, at hourly intervals, and the missing data were padded by the nearest value before or a preset default value. We filtered out 1057 positive and 5834 negative patients in the MIMIC dataset, and 144 positive and 309 negative patients in the HDRJH dataset, respectively. We used a 5-h time window from the patients to predict sepsis. See Appendix 1 for details.

### Machine learning models

In the following two sections, we describe the methodology for developing a sepsis prediction model that outputs the risk of sepsis onset within 5-h preceding at most. To improve the prediction performance in the specific hospital and to avoid the poor performance of direct training due to its insufficient data, the models were first trained in MIMIC and then finetuned in HDRJH using transfer learning techniques. The ultimate sepsis prediction model was obtained by integrating multiple models using ensemble learning techniques.

Multiple models were trained on the MIMIC dataset, including support vector machine (SVM), multilayer perceptron (MLP), gradient boosting machine (GBM), and long short-term memory (LSTM). For GBM, we used XGBoost (26) and LightGBM (27) as implementations. We utilized the standard training methods to train these models with necessary normalization which can be summarized by the following formula:

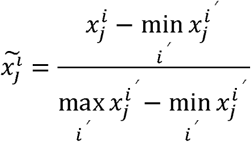

where 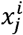 is the value of the j-th feature of the i-th sample, and 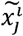 is the value after scalarization. The hyperparameters and structures of each model were tuned based on the validation set.

See Appendix 4 and 5 for details.

### Transfer learning

Based on the integrated considerations of the performance including accuracy, the area under the receiver operating characteristic curve (AUC), sensitivity, and inference speed on the MIMIC dataset, we selected the LightGBM and MLP for sepsis prediction on the RJ Hospital data.

The process of transfer learning can be divided into two steps. First, we normalized the data in the MIMIC dataset and the HDRJH dataset respectively so that the features are all scaled between 0 and 1. Second, we performed the transfer of the model parameters. Specifically, for LightGBM, the previous four hours of features in MIMIC were masked during training to assist transfer learning. After that, the HDRJH dataset was fed to finetune with the initial parameter values taken from the trained model from MIMIC. For MLP, we first froze the parameters of the first three of the six layers of the MIMIC models and initialize the parameters of the last three layers. After training on HDRJH, the models were unfrozen and fine-tuning is performed. We also used an ensemble learning method to integrate the LightGBM and MLP models by taking the inference average. The ensembled model is employed for practical sepsis prediction in RJ Hospital.

### Interpretive analysis

We interpreted our prediction models using Shapley additive explanation (SHAP) (28), a game theory-based approach that assigns an importance value to each feature of each prediction.

### Real-time prediction system

We detail the implementation of our real-time prediction system in (29). When a model inference command is executed, the sepsis early warning module obtains real-time features of the patient from the data integration system via SQL query statements, which are then preprocessed, inferred, and interpreted by the module. The data integration system includes a physical server with the PostgreSQL database for storage of sepsis warning related data and a webserver deploying the portal for user access. The medical device integration hub was placed at the bedside, receiving and transmitting data to the data integration system with a time delay of less than 10 s. Using the network or RS-232 interface, the data integration system can integrate data from IntelliVue Information Center, ventilators, Philips ICCA system, Laboratory Information System (LIS), and Hospital Information System (HIS).

## Results

### Characteristics of patients from different datasets

The baselines of characteristics of patients from MIMIC-III, HDRJH, and Ruijin real-world data were analyzed. As shown in Table S2, the baselines of most characteristics were significantly different between patients from MIMIC-III and patients from HDRJH except for 21 characteristics. Therefore, to avoid the influence of these differences on model performance in the target cohort, retraining of the model was performed on HDRJH using transfer learning technique after training on MIMIC-III.

### Prediction performance on internal and external validation of MIMIC-III

The performance of our sepsis prediction models has been evaluated based on the accuracy, AUC, sensitivity, and specificity on the test set. As shown in Figure S3, the GBM-based models (XGBoost and LightGBM) outperformed others (See Table S3 for the performance of the five models). Furthermore, we compared our LightGBM and MLP models with other proposed models developed from MIMIC-III using Sepsis-3 criteria and reported the prediction outcomes within 5 hours before the onset of sepsis. As shown in Table 1, our LightGBM and MLP models were superior to the others, with AUC of 0.98 and 0.96 respectively. However, it should be noticed that although these models all used MIMIC-III database, there were still differences in the training and test sets due to specific data extraction and sepsis criteria.

**Table 1.** The results of different models on the MIMIC-III dataset.

| Model | Preceding hours | Accuracy | AUC<sup>1</sup> | Sensitivity | Specificity |
| --- | --- | --- | --- | --- | --- |
| InSight | 4 | 0.57 | 0.74 | 0.8 | 0.54 |
| AISE<sup>2</sup> | 4 | 0.64 | 0.84 | 0.85 | 0.64 |
| MGP-TCN<sup>3</sup> | 4 | - | approximately 0.85 | - | - |
| DTW-KNN<sup>4</sup> | 4 | - | approximately 0.88 | - | - |
| MLA<sup>5</sup> | 0 | - | 0.88 | 0.8 | 0.78 |
| MLA | 24 | - | 0.84 | 0.8 | 0.72 |
| DSPA<sup>6</sup> | 4 | - | 0.98 | - | - |
| MGP-AttTCN<sup>7</sup> | 4 | - | 0.75 | - | - |
| NAVOY Sepsis | 3 | 0.81 | 0.84 | 0.74 | 0.83 |
| LightGBM | 4 | 0.91 | 0.98 | 0.85 | 0.97 |
| MLP<sup>8</sup> | 4 | 0.85 | 0.96 | 0.73 | 0.96 |

For the external validation of our LightGBM and MLP models, we evaluated their prediction performances on HDRJH, the dataset from the target medical center (the ICU in RJ Hospital). The AUCs were 0.82–0.86 and 0.84–0.87 respectively on HDRJH from 1–5h preceding, indicating the substantially worsened performance of these models when applied to external independent cohorts.

### Improved prediction performance on HDRJH after transfer learning

To improve the prediction performance on HDRJH, these models were retrained and ensembled using transfer learning and ensemble learning technique. As shown in Table 2, transfer learning improved the prediction performance when deploying the models derived from the public dataset (MIMIC) to the target hospital (HDRJH). The ultimate AUCs of the ensemble sepsis prediction model were 0.94–0.94 from 1–5h preceding on HDRJH, as shown in Table S4.

**Table 2.**
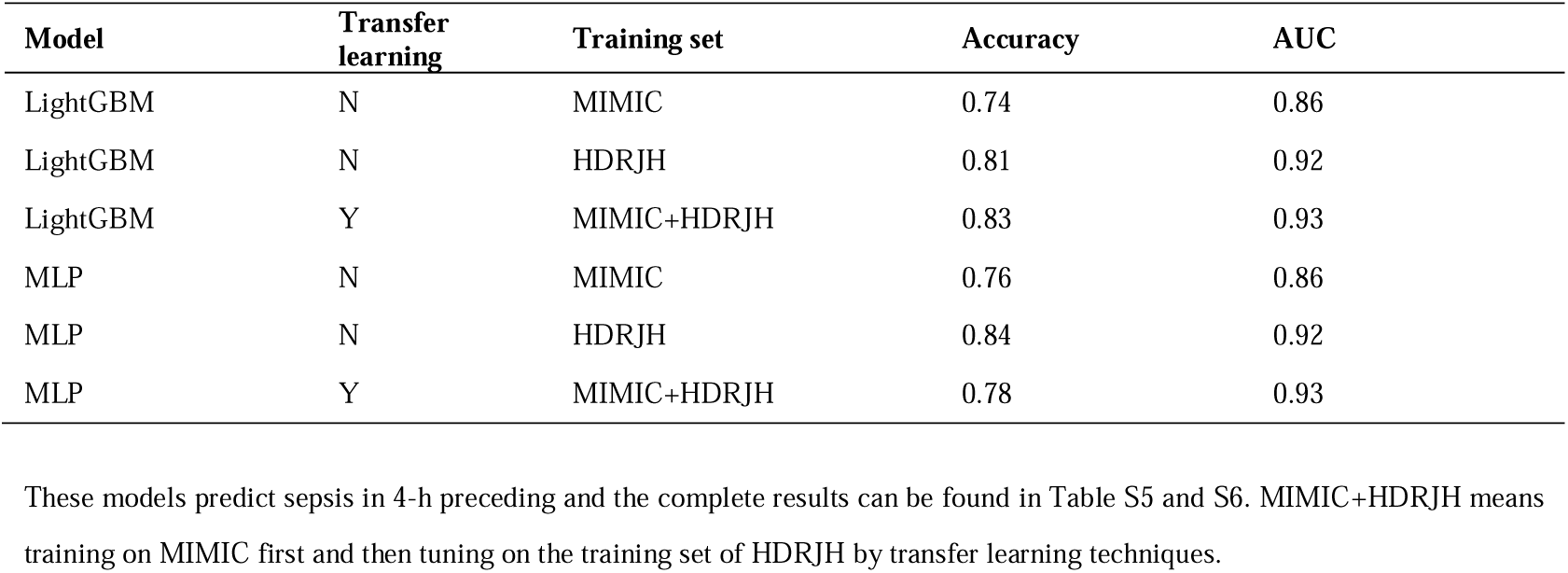
The results of models trained on different datasets on the HDRJH test set.

| Model | Transfer learning | Training set | Accuracy | AUC |
| --- | --- | --- | --- | --- |
| LightGBM | N | MIMIC | 0.74 | 0.86 |
| LightGBM | N | HDRJH | 0.81 | 0.92 |
| LightGBM | Y | MIMIC+HDRJH | 0.83 | 0.93 |
| MLP | N | MIMIC | 0.76 | 0.86 |
| MLP | N | HDRJH | 0.84 | 0.92 |
| MLP | Y | MIMIC+HDRJH | 0.78 | 0.93 |

Furthermore, as shown in Figure 1, LightGBM and MLP models showed consistent transfer benefits on the target hospital at different sampling ratios of the target hospital dataset. Meanwhile, the models after transfer learning showed higher AUCs on MIMIC, indicating improved generalizability of the model to different datasets.

**Figure 1:**
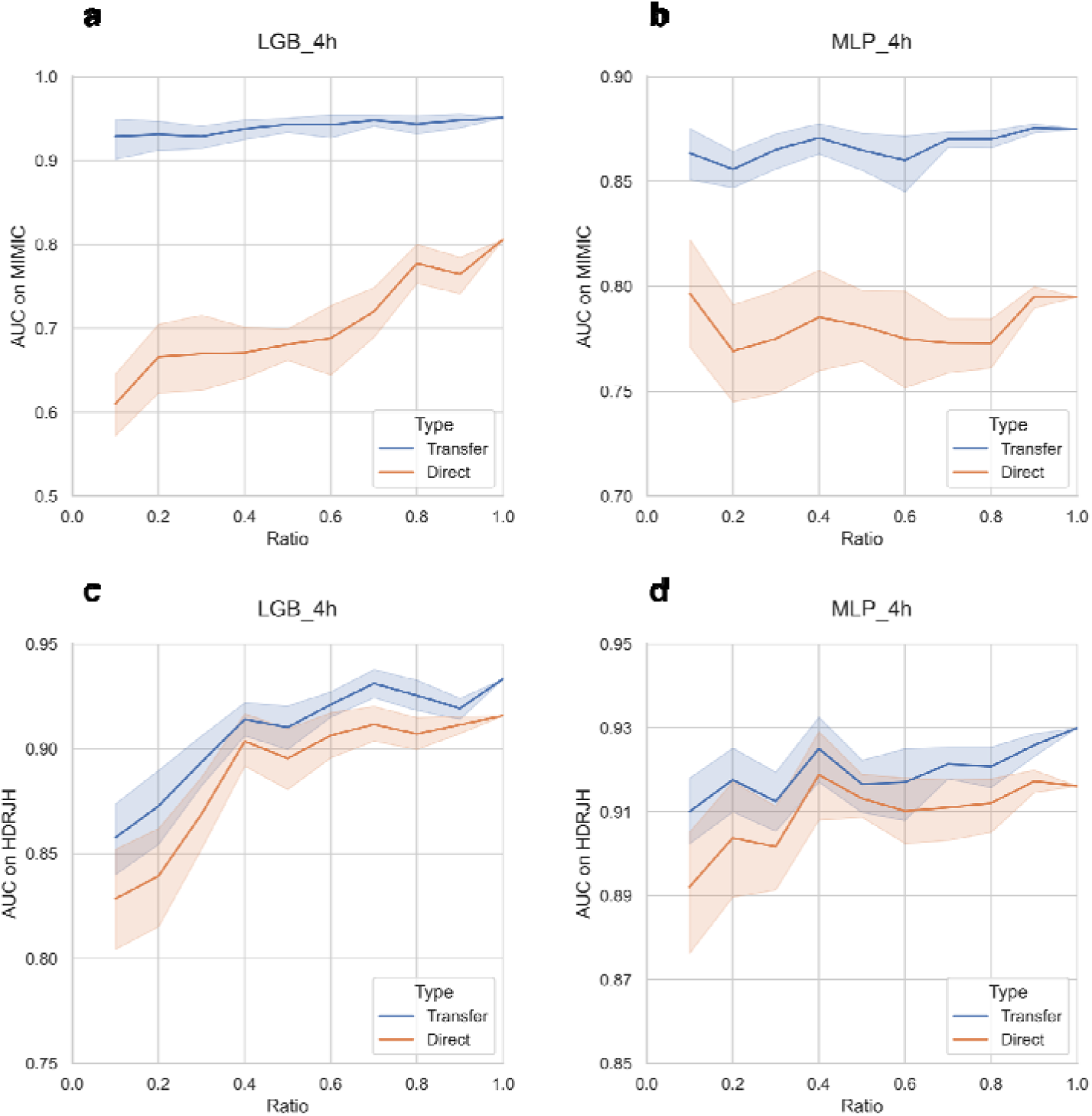
The results of models trained on sampled HDRJH dataset. The training set of HDRJH was sampled at different ratios to simulate a medical center with fewer records. These trained models were tested on the common MIMIC test set (top) and the HDRJH test set (bottom), respectively. The shaded part in the figure represents the 95% confidence interval. These models predict sepsis in 4-h preceding and the complete results can be found in Figure S5-8.

### Feature interpretability of the prediction models

The top 20 features for the LightGBM model predicting sepsis in 4-h preceding were shown in Figure 2, and the results for full analysis of LightGBM and MLP models were shown in Figure S10, 11. Some of these features (antibiotics, respiratory rate, temperature, ventilation, and heart rate) were related to the definition of Sepsis-3 or SIRS. Additionally, the association of some of these features (respiratory rate (30), fibrinogen (31), net balance (32), and age (33)) with the severity or mortality of sepsis has been reported. These data indicate the good interpretability of our prediction model for clinical application.

**Figure 2:**
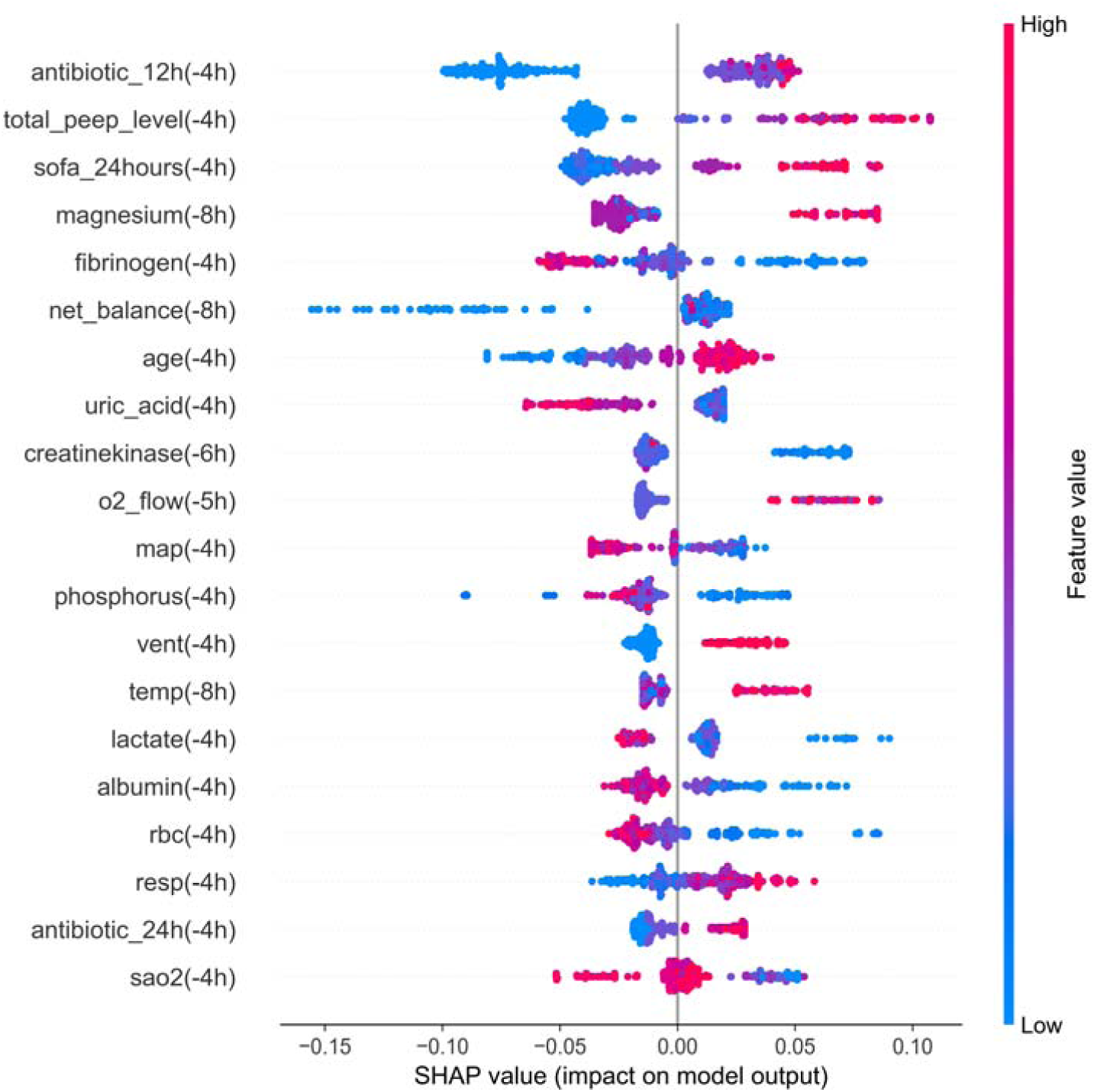
Feature importance of LightGBM model in the sepsis early warning module. The model predicted sepsis in 4-h preceding and the complete results can be found in Figure S10.

### Prediction performance in the real world

For the abovementioned test of model performance, each patient was labeled by the change in SOFA score and the doctor’s examination for infection at a threshold of 0.5. Data from the control group and near onset of sepsis in the case group were included in the analysis. As shown in Table 3, the AUCs for sepsis predictions in 1–5h preceding were 0.86–0.90.

**Table 3.**
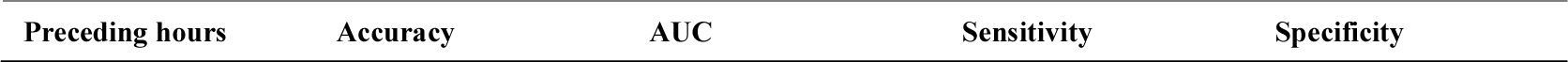
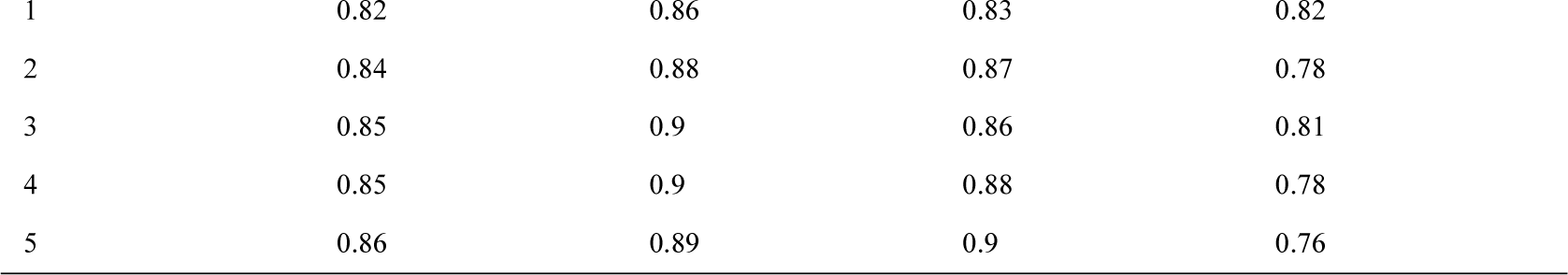
The results of real-world data.

In the real-world study, the threshold was increased to 0.7 to reduce the false alarm rate of sepsis warnings. Figure 3 illustrated examples of the prediction of sepsis by SEPRES over a random period (See Appendix 10 for more details). In the continuously early warning process of 67 patients admitted to the ICU, 22 septic patients and 29 non-septic patients were correctly predicted, whereas 17 non-septic patients and 6 septic patients were incorrectly predicted as false-positive and false-negative cases.

**Figure 3:**
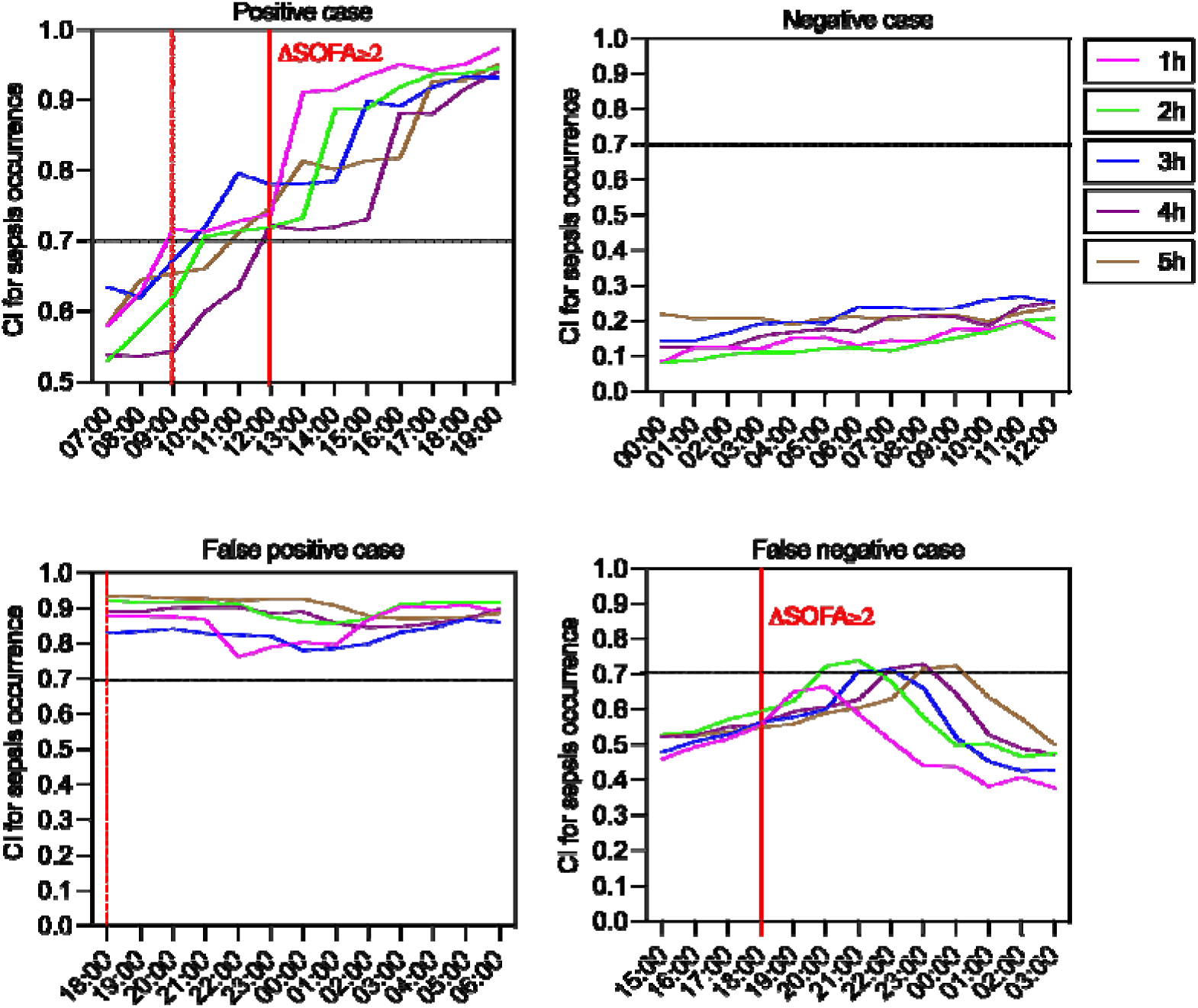
Some illustrative examples of prediction. Each subplot described the confidence index for multiple models (Y-axis) at the indicated time (X-axis). (i) The condition of the patient aggravated in the early morning, with multiple organ dysfunction, and the patient was diagnosed with sepsis at noon. Our model prediction exceeded the warning threshold of 0.7 for the prediction at 9:00 AM. (ii) Despite the high SOFA score (7.0), there was no evidence of ΔSOFA ≥2 within 72 h. Consistently, the predictions were all lower than the threshold. (iii) Although the patient’s SOFA score was stable at 6.0, our model made incorrect predictions of sepsis. (iv) The SOFA score showed an increase from 6.0 to 9.0 at 06:00 PM. In combination with evidence of infection, the patient was diagnosed with sepsis. However, the prediction was below the warning threshold.

## Discussion

Machine learning has been considered a promising method for sepsis prediction in the ICU (6-16,20-22). Early diagnosis and timely management of septic patients can effectively improve the prognosis (34). However, sepsis may not be diagnosed in time in the clinic due to the day-night shift and inattention of medical staff. Therefore, an accurate and efficient early prediction system for sepsis at the bedside is urgently needed. In this study, we established an ICU bedside sepsis early warning system, SEPRES, to conduct real-time sepsis prediction for patients in the ICU by integrating IntelliVue Information Center, ventilators, Philips ICCA system, LIS, and HIS data. Although SEPRES could not provide a definitive basis for our therapeutic regime, the predicted probability of sepsis occurrence allows us to pay more attention to at-sepsis-risk patients.

Generalizability is the major obstacle to the deployment of machine learning into medical practice. Sufficiently large data size is crucial for the training of the machine learning model to achieve good performance. Moreover, the performance of the model derived from one cohort is always worsened when applied to external independent cohorts due to the differences in race, medical environment, disease type, and disease severity in different cohorts. In addition to this, as new tests and techniques are added, new features may help our prediction task, but direct inclusion into existing machine learning models is usually not feasible. In our study, we deployed the transfer learning technique to improve the performance of our models in the target medical center. The transfer learning process effectively improved the prediction AUCs of LightGBM and MLP models on the HDRJH dataset, learns patterns from additional features, and showed consistent benefits across different data sizes of the target cohort. Hence, we argue that transfer learning might be a promising and feasible strategy to maintain the effectiveness of trans-center deployment of machine learning models. Transfer learning can also be used for similar but different prediction tasks, reducing the size requirements of the target dataset, and improving the training speed and the prediction performance (35). In our context, transfer learning can be used to predict different types of diseases, such as disseminated intravascular coagulation (DIC) or acute kidney injury (AKI).

Moreover, the lack of interpretability of these data-driven models prevents the practitioners to trust and accept these machine learning models in the clinic. In the present study, SHAP analysis as the interpretive tool helps medical practitioners identify top risk factors. what needs to be noted is fluid net balance and the use of antibiotics. Due to the difficulty of collecting the net balance data in most datasets, net balance has neither been considered as a feature for most machine learning models based on MIMIC datasets nor been analyzed as an important factor for sepsis prediction inference. Indeed, positive cumulative fluid balance has been reported to be an independent predictor of ICU mortality (32). Moreover, Lin et al have shown that patients with an early positive fluid balance have an increased risk of developing venous thromboembolism (36). Our SHAP analysis further emphasized the importance of careful fluid management in critically ill patients. Therefore, we argue that including the net balance in the prediction model may improve not only the model performance but also the ICU clinical management. In addition, Our SHAP analysis results suggested that antibiotic use tends to increase the predicted occurrence of sepsis. Due to the uncertainty regarding antibiotic initiation in patients with suspected infection, the use of antibiotics is mostly empirical in ICU patients (37). It has been reported that inappropriate antibiotic treatment may accelerate the death of mice via increasing gut proliferation and systemic spreading of a multi-drug resistant (MDR) *Escherichia coli* strain (38). Moreover, the initial inappropriate broad-spectrum antibiotic therapy may promote the dissemination of multidrug-resistant bacteria (MDRB), increase opportunistic infection, and is associated with the poor prognoses of patients (39,40). Therefore, the inappropriate use of antibiotics may aggravate the severity of ICU patients and increase the probability of sepsis. It is urgently needed to improve the management of antibiotic therapy in ICU patients by optimizing reliable specimens and de-escalating antibiotics based on rapid diagnostic tools, therapeutic drug monitoring (TDM), and combined therapy.

SEPRES has certain limitations. First, we enrolled only patients who were non-septic during the entire period in the ICU as negative controls. The enrollment condition may be too pure to establish a model to predict sepsis, which may cause false-positive cases. Second, as we observed in consecutive case studies, patients diagnosed with sepsis shortly after being transferred to the ICU were difficult to be predicted by our model, which is probably due to that our model tends to give lower predictions when the collected data are limited. Finally, variables such as antibiotics and mechanical ventilation were incorporated by our model, resulting in the influence of the model predictions by the subjective behavior of the doctor. However, considering that the use of antibiotics and mechanical ventilation are associated with the severity of the patient, it is essential to include them in our model. These limitations will be addressed in future work through diverse methods, including fine-grained labeling, inclusion of data collection from the ICU, and data augmentation. We also highlight that the application of transfer learning and interpretive tools can significantly improve the generalization and interpretability of the model but still possesses a distance to totally solve it.

## Conclusions

In conclusion, the early prediction of sepsis occurrence by our SEPRES effectively guided medical practitioners to appropriately pay more attention to at-sepsis-risk patients, leading to early diagnosis of sepsis and more efficient ICU patient management. Our SHAP analysis suggests that we can place more emphasis on fluid net balance and the use of antibiotics. Moreover, with the help of the data integration system to collect necessary features and data, the workflow of SEPRES can be applied to disease warnings other than sepsis in the ICU, such as DIC and AKI. Furthermore, the proposed system can be applied to a larger number of medical centers with a certain number of records through transfer learning.

## Data Availability

The data that support the findings of this study are available from the corresponding author upon reasonable request.

## Acknowledgments

The authors acknowledge Shanghai Electric Group Co., Ltd. Central Academe for their support during the development of the data integration system.

## Authors’ contributions

WL, YT, LL, QC, RL, and Lin C conceived and designed the study. YT, LL, QC, RL, DC, HQ, and YH acquired the data. WL, Lin C, and Lai C implemented quality control of data and algorithms. WL, QC, RL, and Lai C had full access to and verified all data in the study. QC developed, trained, and applied machine-learning models. Lai C developed a data integration system. YT, LL, RL, DC, and HQ performed consecutive case studies. QC and RL prepared the first draft of the manuscript. WL, LL, and YT revised the manuscript. All authors contributed to the preparation of the manuscript.

## Availability of supplementary resources

The MIMIC datasets generated and/or analyzed during the current study are available in the MIMIC repository, https://mimic.physionet.org/. The HDRJH dataset and RJ real-world data were approved for limited use by the Ruijin Hospital Ethics Committee, and were not publicly available. The code of the model inference was uploaded into GitHub under “SEPRES”.

## References

1. Cecconi M, Evans L, Levy M, Rhodes A: Sepsis and septic shock. Lancet 2018; 392:75– 87. https://doi.org/10.1016/S0140-6736(18)30696-2

2. Singer M, Deutschman CS, Seymour CW, et al: The third international consensus definitions for sepsis and septic shock (Sepsis-3). Jama 2016; 315:801–810. https://doi.org/10.1001/jama.2016.0287

3. Angus DC, Linde-Zwirble WT, Lidicker J, Clermont G, Carcillo J, Pinsky MR: Epidemiology of severe sepsis in the United States: Analysis of incidence, outcome, and associated costs of care. Crit Care Med 2001; 29:1303–1310. https://doi.org/10.1097/00003246-200107000-00002

4. Marik PE, Farkas JD: The changing paradigm of sepsis: early diagnosis, early antibiotics, early pressors, and early adjuvant treatment. Crit Care Med 2018; 46:1690–1692. https://doi.org/10.1097/CCM.0000000000003310

5. Filbin MR, Lynch J, Gillingham TD, et al: Presenting symptoms independently predict mortality in septic shock: Importance of a previously unmeasured confounder. Crit Care Med 2018; 46:1592–1599. https://doi.org/10.1097/CCM.0000000000003260

6. Henry KE, Hager DN, Pronovost PJ, Saria S: A targeted real-time early warning score (TREWScore) for septic shock. Sci Transl Med 2015; 7:299ra122–299ra122. https://doi.org/10.1126/scitranslmed.aab3719

7. Lauritsen SM, Kalør ME, Kongsgaard EL, et al: Early detection of sepsis utilizing deep learning on electronic health record event sequences. Artif Intell Med 2020; 104:101820. https://doi.org/10.1016/j.artmed.2020.101820

8. Desautels T, Calvert J, Hoffman J, et al: Prediction of sepsis in the intensive care unit with minimal electronic health record data: A machine learning approach. JMIR Med Inform 2016; 4:e5909. https://doi.org/10.2196/medinform.5909

9. Nemati S, Holder A, Razmi F, Stanley MD, Clifford GD, Buchman TG: An interpretable machine learning model for accurate prediction of sepsis in the ICU. Crit Care Med 2018; 46:547. https://doi.org/10.1097/CCM.0000000000002936

10. Moor M, Horn M, Rieck B, Roqueiro D, Borgwardt K: Early recognition of sepsis with Gaussian process temporal convolutional networks and dynamic time warping. Machine Learning for Healthcare Conference, PMLR 2019; 106:2–26

11. Barton C, Chettipally U, Zhou Y, et al: Evaluation of a machine learning algorithm for up to 48-hour advance prediction of sepsis using six vital signs. Comput Biol Med 2019; 109:79–84. https://doi.org/10.1016/j.compbiomed.2019.04.027

12. Asuroglu T, Ogul H: A deep learning approach for sepsis monitoring via severity score estimation. Comput Methods Programs Biomed 2021; 198:105816. https://doi.org/10.1016/j.cmpb.2020.105816

13. Rosnati M, Fortuin V: MGP-AttTCN: An interpretable machine learning model for the prediction of sepsis. PLoS One 2021; 16:e0251248. https://doi.org/10.1371/journal.pone.0251248

14. Persson I, Östling A, Arlbrandt M, Söderberg J, Becedas D: A Machine Learning Sepsis Prediction Algorithm for Intended Intensive Care Unit Use (NAVOY Sepsis): Proof-of-Concept Study. JMIR Form Res 2021; 5(9):e28000. https://doi.org/10.2196/28000

15. McCoy A, Das R: Reducing patient mortality, length of stay and readmissions through machine learning-based sepsis prediction in the emergency department, intensive care unit and hospital floor units. BMJ Open Qual 2017; 6:e000158. https://doi.org/10.1136/bmjoq-2017-000158

16. Shimabukuro DW, Barton CW, Feldman MD, Mataraso SJ, Das R: Effect of a machine learning-based severe sepsis prediction algorithm on patient survival and hospital length of stay: A randomised clinical trial. BMJ Open Respir Res 2017; 4:e000234. https://doi.org/10.1136/bmjresp-2017-000234

17. Cho K-J, Kwon O, Kwon J-m, Lee Y, Park H, Jeon K-H, et al: Detecting patient deterioration using artificial intelligence in a rapid response system. Crit Care Med 2020; 48(4):e285–e9. https://doi.org/10.1097/ccm.0000000000004236

18. Siontis GC, Tzoulaki I, Castaldi PJ, Ioannidis JP: External validation of new risk prediction models is infrequent and reveals worse prognostic discrimination. J Clin Epidemiol 2015; 68(1):25–34. https://doi.org/10.1016/j.jclinepi.2014.09.007

19. Lee J, Maslove DM: Customization of a severity of illness score using local electronic medical record data. J Intensive Care Med 2017; 32(1):38–47. https://doi.org/10.1177/0885066615585951

20. Mao Q, Jay M, Hoffman JL, Calvert J, Barton C, Shimabukuro D, et al: Multicentre validation of a sepsis prediction algorithm using only vital sign data in the emergency department, general ward and ICU. BMJ open 2018; 8(1):e017833. https://doi.org/10.1136/bmjopen-2017-017833

21. Wardi G, Carlile M, Holder A, Shashikumar S, Hayden SR, Nemati S: Predicting progression to septic shock in the emergency department using an externally generalizable machine-learning algorithm. Ann Emerg Med 2021; 77(4):395–406. https://doi.org/10.1016/j.annemergmed.2020.11.007

22. Zhang D, Yin C, Hunold KM, Jiang X, Caterino JM, Zhang P: An interpretable deep-learning model for early prediction of sepsis in the emergency department. Patterns (N Y) 2021; 2:100196. https://doi.org/10.1016/j.patter.2020.100196

23. Gandin I, Scagnetto A, Romani S, Barbati G: Interpretability of time-series deep learning models: A study in cardiovascular patients admitted to Intensive care unit. J Biomed Inform 2021; 121:103876. https://doi.org/10.1016/j.jbi.2021.103876

24. Collins GS, Reitsma JB, Altman DG, Moons KG: Transparent reporting of a multivariable prediction model for individual prognosis or diagnosis (TRIPOD): the TRIPOD statement. Ann Intern Med 2015; 162:55–63. https://doi.org/10.1136/bmj.g7594

25. Johnson AE, Pollard TJ, Shen L, et al: MIMIC-III, a freely accessible critical care database. Sci Data 2016; 3:1–9. https://doi.org/10.1038/sdata.2016.35

26. Chen T, Guestrin C: Xgboost: A scalable tree boosting system. Proceedings of the 22nd acm sigkdd international conference on knowledge discovery and data mining 2016; 785–794. https://doi.org/10.1145/2939672.2939785

27. Ke G, Meng Q, Finley T, et al: Lightgbm: A highly efficient gradient boosting decision tree. Advances in neural information processing systems 2017; 30:3146–3154

28. Lundberg S, Lee SI: A Unified approach to interpreting model predictions. Proceedings of the 31st international conference on neural information processing systems 2017; 4768–4777

29. Chen Q, Li R, Lin C, et al: SEPRES: Sepsis prediction via the clinical data integration system in the ICU. medRxiv 2022

30. Kenzaka T, Okayama M, Kuroki S, et al: Importance of vital signs to the early diagnosis and severity of sepsis: Association between vital signs and sequential organ failure assessment score in patients with sepsis. Intern Med 2012; 51:871–876. https://doi.org/10.2169/internalmedicine.51.6951

31. Matsubara T, Yamakawa K, Umemura Y, et al: Significance of plasma fibrinogen level and antithrombin activity in sepsis: A multicenter cohort study using a cubic spline model. Thromb Res 2019; 181:17–23. https://doi.org/10.1016/j.thromres.2019.07.002

32. Brotfain E, Koyfman L, Toledano R, et al: Positive fluid balance as a major predictor of clinical outcome of patients with sepsis/septic shock after ICU discharge. Am J Emerg Med 2016; 34:2122–2126. https://doi.org/10.1016/j.ajem.2016.07.058

33. Martin GS, Mannino DM, Moss M: The effect of age on the development and outcome of adult sepsis. Crit Care Med 2006; 34:15–21. https://doi.org/10.1097/01.CCM.0000194535.82812.BA

34. Burdick H, Pino E, Gabel-Comeau D, et al: Effect of a sepsis prediction algorithm on patient mortality, length of stay and readmission: A prospective multicentre clinical outcomes evaluation of real-world patient data from US hospitals. BMJ Health Care Inform 2020; 27:e100109. https://doi.org/10.1136/bmjhci-2019-100109

35. Morid MA, Borjali A, Del Fiol G: A scoping review of transfer learning research on medical image analysis using ImageNet. Comput Biol Med 2021; 128:104115. https://doi.org/10.1016/j.compbiomed.2020.104115

36. Lin T-L, Dhillon NK, Conde G, et al: Early positive fluid balance is predictive for venous thromboembolism in critically ill surgical patients. Am J Surg 2021; 222:220–226. https://doi.org/10.1016/j.amjsurg.2020.08.032

37. Klein Klouwenberg PM, Cremer OL, van Vught LA, et al: Likelihood of infection in patients with presumed sepsis at the time of intensive care unit admission: a cohort study. Crit Care 2015; 19:319. https://doi.org/10.1186/s13054-015-1035-1

38. Ayres JS, Trinidad NJ, Vance RE: Lethal inflammasome activation by a multidrug-resistant pathobiont upon antibiotic disruption of the microbiota. Nat Med 2012; 18:799– 806. https://doi.org/10.1038/nm.2729

39. Timsit JF, Bassetti M, Cremer O, et al: Rationalizing antimicrobial therapy in the ICU: a narrative review. Intensive Care Med 2019; 45:172–189. https://doi.org/10.1007/s00134-019-05520-5

40. Luyt CE, Bréchot N, Trouillet JL, Chastre J: Antibiotic stewardship in the intensive care unit. Crit Care 2014; 18:480. https://doi.org/10.1186/s13054-014-0480-6

