## supplementary material for "Transferability and interpretability of the sepsis prediction models in the intensive care unit"

### Appendix 1 Details of data acquisition

***Inclusion criteria***

Patients who met all the following criteria were included in the case group:

1) At least 14 years old.

2) Sepsis onset at least 5 h after admission to the ICU.

3) Sepsis onset is the first instance since admission to the hospital.

Patients who met all the following criteria were included in the control group:

1) At least 14 years old.

2) Patients who stayed in the ICU for at least 5 h and had no sepsis at this stay.

3) Patients without ICD-9 codes for sepsis (785.52, 995.91, and 995.92).

4) SOFA score changes of no more than 1 point in an arbitrary continuous 72 h in the ICU stay.

The third criterion was excluded for the control cases in the HDRJH because ICD-9 codes were not recorded.

***Sepsis definitions***

Patients were followed up throughout their stay in the ICU until discharge or development of sepsis according to the definition of the Third International Consensus for sepsis (Sepsis-3) [2]. Specifically, if the timestamp of antibiotics (*t_abx_*) and blood cultures (*t_culture_*) meet the condition $t_{abx}-24 h\leq t_{culture}\leq t_{abx}+72 h$, the earlier timestamp of *t_abx_* and *t_culture_* is defined as the timestamp of suspected infection (*t_sus_*). The SOFA score was evaluated per hour within the time window $\left[ t_{sus}-48 h,t_{sus}+24 h \right]$. The first hour with two or more points of increase in the SOFA score than the lowest prior score is defined as the onset of sepsis (*t_onset_*).

***Feature extraction***

Seventy-eight patient variables from MIMIC were chosen as raw data for the dataset. Appendix 2 contains a complete list of these variables. We excluded significantly incorrect records by setting the range of variables according to the specialists. When integrating the same variables from different sources, we set priorities to extract values with the highest confidence. After data cleaning, these data were summarized per hour into the maximum, average, median, and minimum, except for some changeless or durative variables, which in total were 285 features. Padding was used if there was no value at the corresponding time. Padding values were taken as the nearest value before, or the average of all patients when no value was valid since the patient’s admission. Patients with too few valid variables were removed to ensure data quality. We used a five-hour-long time window from the patients to predict sepsis; thus, each sample point in these tasks had 1425 features. Finally, we obtained a dataset with 1057 positive and 5834 negative patients, which is shown in Figure S1. We divided the dataset into training, validation, and test sets. Negative patients were divided at a ratio of 7:1:2. For positive patients, we chose the same number as the negative patients in the validation and test sets. The remaining positive patients were included in the training set. Oversampling of positive sample points or down-sampling of negative sample points was used to ensure that the proportion was 1:1 in each set.

Similar preparation steps were used in the HDRJH as well, although only sixty-three variables were available. Appendix 3 contains a complete list of these variables. These variables were summarized hourly into 226 features. After padding and filtering, we obtained the HDRJH dataset with 144 positive and 309 negative patients, which is shown in Figure S2. We divided them into a training set with 102 positive and 267 negative patients, a validation set with 12 positive and 12 negative patients, and a test set with 30 positive and 30 negative patients.

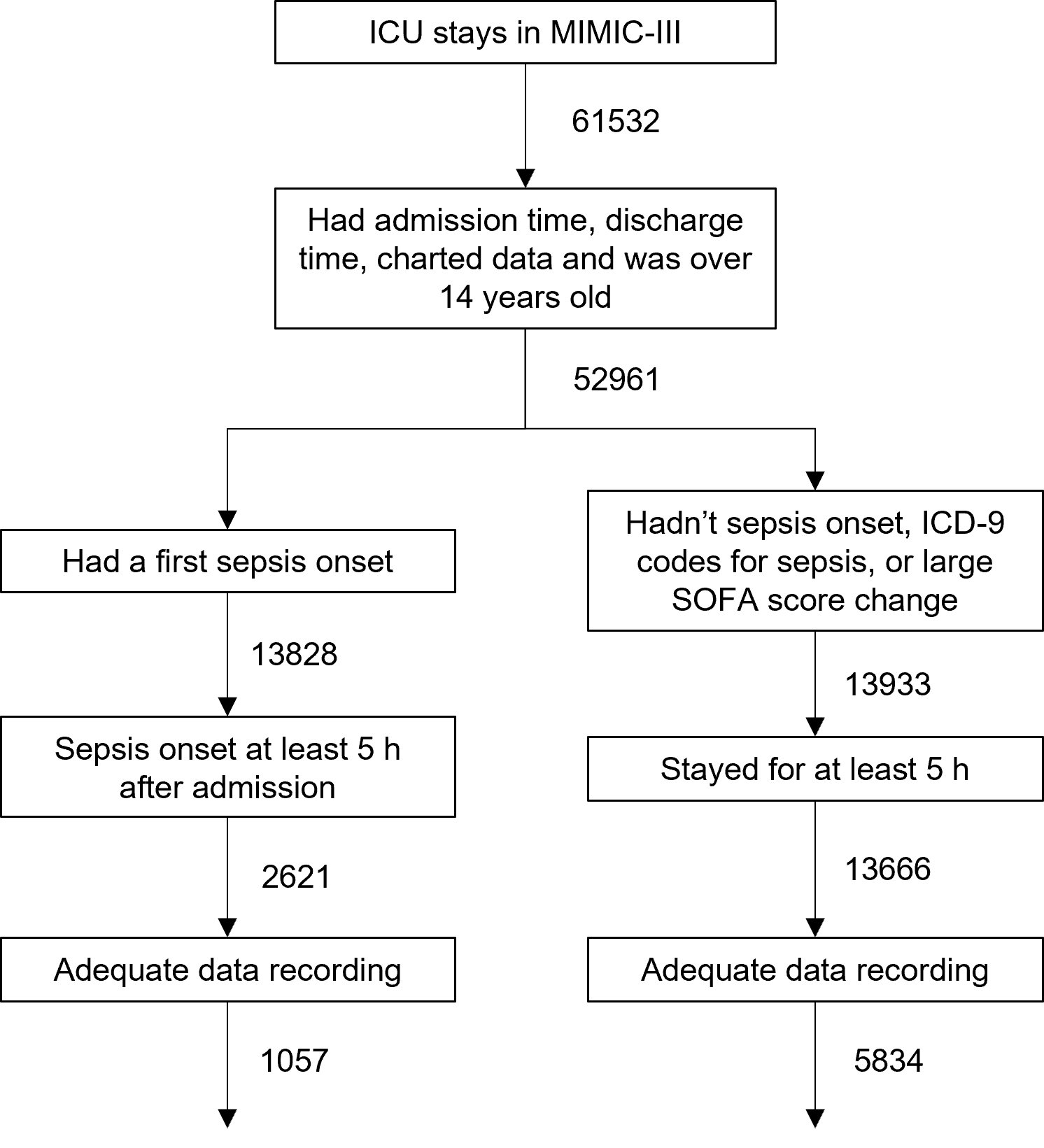

Figure S1. **Flowchart of patient inclusion in MIMIC.**

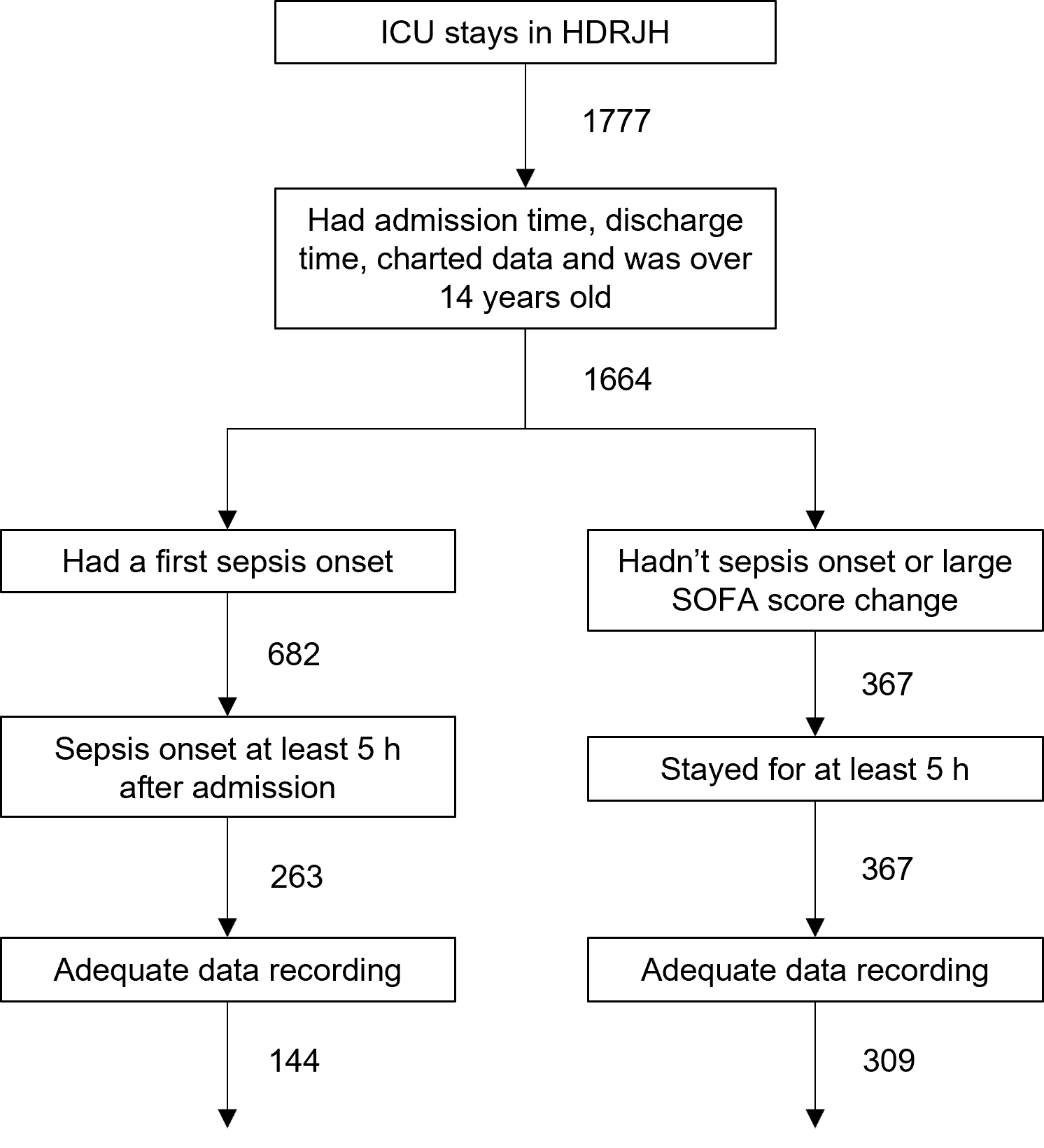

Figure S2. **Flowchart of patient inclusion in HDRJH.**

### Appendix 2 Complete list of variables used on the MIMIC dataset

We consulted literature on predicting sepsis or SOFA scores and the variables mentioned in the literature and could be extracted from MIMIC-III were selected, for a total of 78 variables [1-4].

The 78 variables are MAP, heart rate, O_2_sat, SBP, DBP, respiratory rate, temperature, GCS, PaO_2_, FiO_2_, SpO_2_, cardiac output, stroke volume, stroke volume variation, tidal volume, peak inspiratory pressure, total PEEP level, O_2_ flow rate, WBC, hemoglobin, hematocrit, creatinine, bilirubin, bilirubin direct, platelets, INR, PTT, AST, lactate, glucose, potassium, calcium, BUN, phosphorus, magnesium, chloride, BNP, troponin I, fibrinogen, CRP, sedimentation rate, ammonia, PH, PCO_2_, bicarbonate, base excess, SaO_2_, anion gap, albumin, bands, PT, sodium, ferritin, transferrin, creatine kinase, creatine kinase-MB, LDH, troponin T, RDW, ALP, MCHC, uric acid, monocytes, lymphocytes, MCH, AaDO_2_, RBC, MCV, neutrophils, weight, urine output in the past 24 hours, net balance, ventilation, number of antibiotics in the past 12, 24, and 48 hours, SOFA, and age.

### Appendix 3 Complete list of variables used on the HDRJH dataset

Based on Appendix 2, we removed some variables that were not recorded in the HDRJH and removed some infrequently used variables based on doctors’ opinions. We also added four variables that were involved in the SOFA score.

Nineteen variables were removed: GCS, O_2_sat, Cardiac Output, Stroke Volume, Stroke Volume Variation, Calcium, BNP, CRP, Sedimentation Rate, Ammonia, Anion Gap, Bands, Ferritin, Transferrin, Troponin T, RDW, MCHC, MCH, MCV.

Four variables were added: rate of norepinephrine, epinephrine, dopamine, and dobutamine.

Finally, 63 variables were collected.

### Appendix 4 Machine-learning models

1. Support Vector Machine (SVM)

The rationale of Support Vector Machine (SVM) is to find such a hyperplane
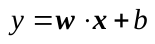
 to separate data that the distance between positive data on one side and the hyperplane and the distance between negative data on the other side and the hyperplane is maximized:

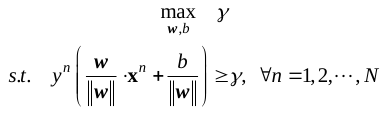

Where the dataset is
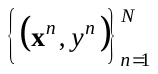
, and
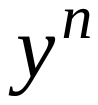
 is the label of
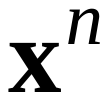
.

Furthermore, kernel tricks can be introduced to improve the nonlinearity of the model. Data are mapped into a feature space using a nonlinear mapping with the help of kernel function, including polynomial kernel function, linear kernel, or mixed kernel function.

2. Multi-Layer Perceptron (MLP)

Multi-Layer Perceptron (MLP) is a classical and useful neural network model. It contains an input layer, several hidden layers, and an output layer.

Let *In* denote the input layer, and
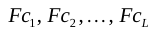
 denote hidden layers and the output layer successively, where
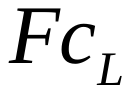
 is the output layer. Each hidden layer and output layer learn a nonlinear map
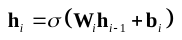
, where
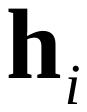
 is the representation of an example at layer
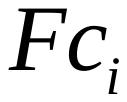
,
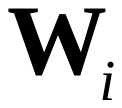
 and
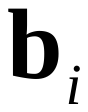
 are the weight and bias parameters of layer
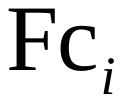
, and
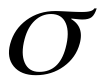
 is the activation function, taken as Rectifier Linear Units (ReLu)
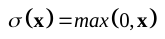
 for the hidden layers and Softmax Units
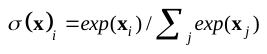
 for the output layer. The loss function like

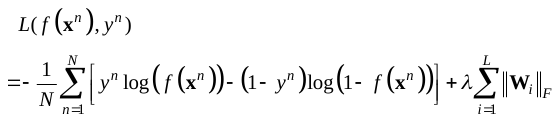

and Stochastic Gradient Descent algorithms are always used to train the networks, where
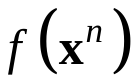
 is the network output of
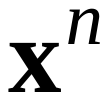
, and
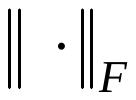
 is the Frobenius norm. The first term in the formula is the cross-entropy loss and the second is a weight decay term in order to prevent over-fitting where
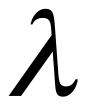
 is the weight decay coefficient.

3. Gradient Boosting Decision Tree

Gradient boosting decision tree (GBDT) is a widely-used machine learning algorithm and there are many effective implementations such as XGBoost and LightGBM which have been used in our experiments. XGBoost makes use of second-order information of loss function instead of first-order information in the case of GBDT, which makes convergence faster and better. Moreover, XGBoost uses several algorithms and technologies to accelerate the training procedure. Compared with XGBoost, LightGBM can accelerate the training further, benefiting from the histogram-based algorithm, gradient-based one-side sampling, and exclusive feature bundling.

4. Long Short-Term Memory (LSTM)

Considering the time-sequential datasets, recurrent neural networks (RNN), specifically long short-term memory (LSTM) networks, which is the most successful RNN, had been used in our experiments. LSTM networks have memory blocks consisting of memory cells and gates in the recurrent hidden layer. The gates control which information to forget and which to remember. Through these, LSTM networks are able to store information selectively and capture the long-time-dependent behavior compared with MLP or GBDT. A single repeating structure of the LSTM can be represented as the following:

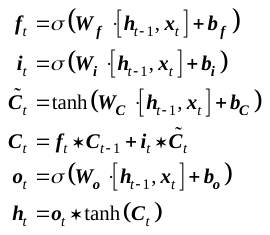

Where
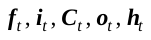
 represent the forget gate, input gate, cell state, output gate, hidden state, and the symbol
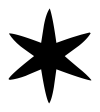
 refers to element-wise multiplication.

### Appendix 5 Training method

Some redundant features were removed to accelerate the training of the SVM and MLP on the MIMIC dataset. For SVM, all average features and features whose coefficient of variation was > 2 were used; for MLP, only one of the maximum, average, median, and minimum of each laboratory variable was kept, considering the low record frequency. Data were standardized (i.e., each feature’s value range was linearly scaled between 0 and 1) before training to eliminate magnitude differences among features and reduce distribution differences between the two datasets in the next task of transfer learning.

We chose the linear kernel function and 1 as the penalty factor in the SVM. For MLP, 460 features were selected as the input. We finally used a six-layer architecture, which is shown in Table S1. We chose 256 as the batch size, 0.001 as the learning rate, 0.6 as the dropout rate, and 0.001 as the weight decay coefficient in MLP. We also used dropout to prevent over-fitting as well. The proposed networks were trained with the AdaGrad algorithm. For XGBoost, we set max_depth as 6, colsample_bytree as 0.2, and other configurations as default. For LightGBM, we set num_leaves as 5, lambda_l2 as 0.1, learning rate as 0.2, and other configurations as default. During LSTM training, we used LSTM architecture with four hidden LSTM layers with 16 one-cell memory blocks and a fully connected layer with one output unit added, followed by a sigmoid function. We set the learning rate as 0.0001 and the batch size to 2000.

Table S1. **The network structure of the MLP**

|  | **input** | **layer1** | **layer2** | **layer3** | **layer4** | **layer5** | **output** |
| --- | --- | --- | --- | --- | --- | --- | --- |
| **units** | 460 | 256 | 128 | 64 | 24 | 24 | 2 |

### Appendix 6 Characteristics of patients from different datasets

Table S2. **Variables of the three data sources on admission**

| **Variable^[1]^** | **MIMIC (n=6891)** | | **HDRJH (n=453)** | | **RJ real-world (n=67)** | |
| --- | --- | --- | --- | --- | --- | --- |
|  | **Number (%)** | **Missing** | **Number (%)** | **Missing** | **Number (%)** | **Missing** |
| Gender-male | 3995 (57.97) | 0 | 258 (56.95) | 0 | 43 (65.15) | 1 |
| Ventilation | 2166 (31.43) | 0 | 172 (37.97) | 0 | 33 (50) | 1 |
|  | **Mean (SD)** | **Missing** | **Mean (SD)** | **Missing** | **Mean (SD)** | **Missing** |
| SOFA ^**^ | 2.67 (2.37) | 0 | 3.13 (2.53) | 0 | 4.64 (3.37) | 0 |
| Age (year) | 61.32 (17.15) | 0 | 61.76 (18.66) | 4 | 63.36 (17.13) | 1 |
| Weight (kg) ^***^ | 83.18 (24.38) | 299 | 63.54 (15.38) | 147 | 63.54 (14.29) | 3 |
| Heart rate (bpm) ^**^ | 84.25 (17.3) | 4 | 88.18 (18.65) | 0 | 88.83 (23.33) | 2 |
| Systolic blood pressure (mmHg) ^***^ | 123.85 (22.5) | 4 | 132.15 (20.97) | 2 | 130.75 (27.63) | 2 |
| Diastolic blood pressure (mmHg) ^***^ | 63.75 (15.23) | 4 | 75.04 (14.46) | 2 | 75.92 (18.54) | 2 |
| Mean arterial pressure (mmHg) ^***^ | 81.35 (16.47) | 4 | 88.34 (14.67) | 2 | 93.18 (17.91) | 2 |
| Respiratory rate (insp/min) | 18.93 (5.37) | 6 | 19.42 (4.76) | 0 | 19.74 (5.34) | 2 |
| Temperature (Celsius) ^***^ | 36.85 (0.72) | 8 | 37.29 (0.62) | 0 | 37.03 (0.79) | 2 |
| SpO2 (%) ^***^ | 97.09 (3.04) | 4 | 99.12 (1.91) | 0 | 98.44 (4.31) | 19 |
| pH | 7.39 (0.07) | 3006 | 7.4 (0.05) | 1 | 7.4 (0.07) | 2 |
| PaO2 (mmHg) ^***^ | 142.84 (77.85) | 3486 | 108.68 (42.29) | 53 | 139.73 (65.91) | 2 |
| SaO2 (%) | 96.41 (4.14) | 5084 | 96.6 (5.55) | 1 | 99.04 (1.27) | 2 |
| AaDO2 (mmHg) ^***^ | 461.85 (121.42) | 6203 | 110.02 (83.84) | 12 | 129.86 (84.33) | 2 |
| PCO2 (mmHg) ^**^ | 41.69 (9.65) | 3378 | 39.49 (8.74) | 1 | 41.09 (14.31) | 2 |
| Bicarbonate (mEq/L) ^**^ | 24.93 (4.42) | 3 | 24.01 (3.12) | 1 | 23.94 (5.31) | 2 |
| Base Excess | 0.29 (4.77) | 3326 | -0.22 (3.68) | 1 | -0.13 (5.52) | 2 |
| White blood cell count (10^12^/L) | 11.32 (8.93) | 1 | 10.75 (4.78) | 0 | 10.32 (6) | 2 |
| Neutrophils (%) | 78.81 (13.73) | 3801 | 80.9 (12.18) | 3 | 85.85 (12.78) | 2 |
| Monocytes (%) | 4.7 (3.4) | 3801 | 4.55 (4.77) | 4 | 3.43 (2.68) | 2 |
| Lymphocytes (%) | 12.97 (10.6) | 3801 | 12.23 (9.22) | 0 | 8.31 (5.24) | 2 |
| Red blood cell count (10^12^/L) | 3.61 (0.64) | 2 | 3.55 (0.76) | 0 | 5.1 (15.13) | 2 |
| Hemoglobin (g/dL) ^*^ | 10.85 (1.9) | 1 | 10.43 (2.23) | 0 | 9.74 (2.55) | 3 |
| Hematocrit (%) | 32.06 (5.39) | 0 | 31.31 (6.52) | 0 | 30.97 (7.68) | 3 |
| Platelets (10^12^/L) ^*^ | 238.27 (129.13) | 1 | 217.08 (111.56) | 0 | 180.02 (118.32) | 2 |
| BUN (mg/dL) ^***^ | 24.52 (21.25) | 1 | 6.63 (4.7) | 260 | 9.99 (8.38) | 2 |
| Creatinine (mg/dL) ^***^ | 1.35 (1.64) | 1 | 0.96 (1.04) | 2 | 1.7 (2.32) | 2 |
| Uric acid (mg/dL) ^***^ | 5.68 (3.01) | 6594 | 3.98 (2.1) | 13 | 4.87 (2.57) | 2 |
| LDH (IU/L) | 394.09 (806.44) | 4057 | 250.06 (162.55) | 232 | 505.29 (1849.89) | 2 |
| ALP (U/L) ^*^ | 115.37 (124.08) | 2946 | 91.89 (93.57) | 3 | 92.48 (59.21) | 2 |
| AST (U/L) | 131.16 (562.35) | 2857 | 56.17 (152.79) | 2 | 177.83 (851.23) | 2 |
| Bilirubin (mg/dL) | 1.61 (4.07) | 2889 | 1.64 (2.42) | 2 | 1.61 (1.65) | 2 |
| Bilirubin direct (mg/dL) ^***^ | 3.36 (4.94) | 6387 | 10.8 (23.92) | 6 | 10.98 (17.33) | 2 |
| Albumin (g/dL) | 3.11 (0.65) | 3637 | 3.09 (0.5) | 1 | 3.03 (0.51) | 2 |
| Partial thromboplastin time (sec) | 36.08 (19.9) | 616 | 35.71 (17.02) | 3 | 32.89 (13.33) | 2 |
| Prothrombin time (sec) ^*^ | 15.04 (5.86) | 594 | 14.04 (4.54) | 3 | 14.72 (3.53) | 2 |
| INR ^***^ | 1.38 (0.71) | 594 | 1.18 (0.27) | 3 | 1.26 (0.32) | 2 |
| Fibrinogen (mg/dL) ^***^ | 365.79 (199.95) | 5711 | 292.56 (131.82) | 8 | 339.38 (120.92) | 2 |
| Lactate (mmol/L) ^***^ | 1.85 (1.5) | 4091 | 2.48 (1.92) | 51 | 2.25 (1.64) | 3 |
| Glucose (mg/dL) ^***^ | 135.77 (55.62) | 0 | 156.29 (54.24) | 69 | 193.65 (87.6) | 2 |
| Sodium (mEq/L) | 138.64 (4.42) | 1 | 138.81 (5.8) | 113 | 138.98 (6.7) | 2 |
| Chloride (mEq/L) ^***^ | 104.64 (5.64) | 2 | 101.89 (4.98) | 0 | 106.32 (6.35) | 2 |
| Potassium (mEq/L) ^***^ | 4.07 (0.6) | 0 | 3.86 (0.45) | 2 | 3.92 (0.63) | 2 |
| Phosphorus (mEq/L) ^***^ | 3.5 (1.25) | 306 | 3.05 (1.18) | 54 | 3.37 (1.76) | 2 |
| Magnesium (mg/dL) | 2.01 (0.36) | 80 | 2 (0.35) | 173 | 2.05 (0.39) | 16 |
| Troponin I (ng/mL) ^***^ | 6.14 (9.28) | 6656 | 0.26 (2.49) | 235 | 0.25 (0.68) | 7 |
| Creatine Kinase (IU/L) | 716.22 (3592.92) | 3314 | 258.64 (895.02) | 232 | 175.38 (254.81) | 1 |
| Creatine Kinase MB (ng/mL) ^***^ | 30.53 (73.44) | 4075 | 2.93 (4.45) | 234 | 3.8 (6.12) | 1 |

### Appendix 7 Results of sepsis prediction models on the MIMIC dataset

Table S3. **The results of models on the MIMIC dataset**

|  | **Preceding hours** | Accuracy | AUC | Sensitivity | Specificity |
| --- | --- | --- | --- | --- | --- |
| **SVM** | 1 | 0.88 | 0.88 | 0.87 | 0.88 |
|  | 2 | 0.87 | 0.87 | 0.87 | 0.87 |
|  | 3 | 0.88 | 0.88 | 0.88 | 0.88 |
|  | 4 | 0.87 | 0.87 | 0.87 | 0.86 |
|  | 5 | 0.87 | 0.87 | 0.87 | 0.87 |
| **MLP** | 1 | 0.84 | 0.95 | 0.71 | 0.98 |
|  | 2 | 0.84 | 0.95 | 0.71 | 0.97 |
|  | 3 | 0.84 | 0.96 | 0.71 | 0.97 |
|  | 4 | 0.85 | 0.96 | 0.73 | 0.96 |
|  | 5 | 0.85 | 0.96 | 0.74 | 0.96 |
| **XGBoost** | 1 | 0.89 | 0.99 | 0.8 | 0.98 |
|  | 2 | 0.9 | 0.99 | 0.81 | 0.98 |
|  | 3 | 0.9 | 0.99 | 0.81 | 0.98 |
|  | 4 | 0.9 | 0.99 | 0.82 | 0.98 |
|  | 5 | 0.9 | 0.99 | 0.82 | 0.98 |
| **LightGBM** | 1 | 0.89 | 0.98 | 0.81 | 0.97 |
|  | 2 | 0.9 | 0.98 | 0.82 | 0.97 |
|  | 3 | 0.9 | 0.98 | 0.83 | 0.96 |
|  | 4 | 0.91 | 0.98 | 0.85 | 0.97 |
|  | 5 | 0.91 | 0.98 | 0.84 | 0.97 |
| **LSTM** | 1 | 0.84 | 0.95 | 0.7 | 0.97 |
|  | 2 | 0.85 | 0.95 | 0.74 | 0.96 |
|  | 3 | 0.86 | 0.95 | 0.77 | 0.95 |
|  | 4 | 0.85 | 0.95 | 0.76 | 0.94 |
|  | 5 | 0.87 | 0.95 | 0.82 | 0.92 |

Table S3 shows that the AUCs of XGBoost and LightGBM were the highest among these sepsis prediction models, followed by MLP and LSTM, and SVM performed the worst. Although the GBM’s structure was relatively simple, it outperformed the artificial neural network models, which may be due to the complexity of artificial neural network models, leading to poor generalization. At the same time, LSTM didn’t outperform MLP. In each model, the five tasks of predicting sepsis from 1 to 5 h in advance did not show significant differences in the predicting AUC.

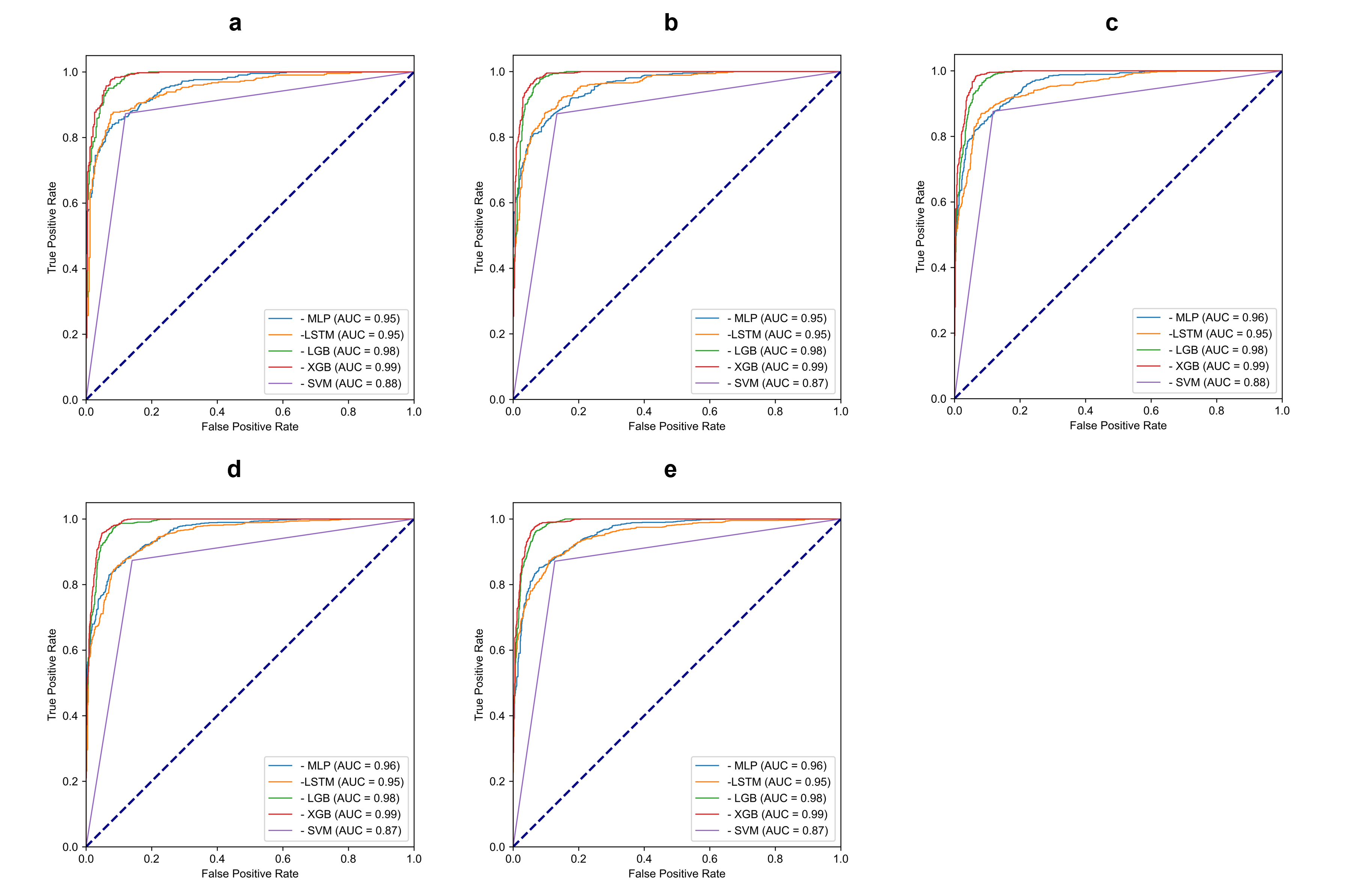

Figure S3. **ROC curves of models detecting sepsis in 1–5-h preceding on the MIMIC-III dataset.** The AUCs of XGBoost and LightGBM were the highest among these sepsis prediction models, followed by MLP and LSTM, and SVM.

### Appendix 8 Results of sepsis prediction models on the HDRJH dataset

The ultimate performances of the ensemble sepsis prediction model are shown in Table S4, and The complete performances of LightGBM and MLP models trained on different datasets on the HDRJH test set are shown in Table S5 and Table S6, where MIMIC+ HDRJH means training on MIMIC first, and then tuning on the training set of HDRJH by transfer learning technique. Models trained using the transfer learning technique performed the best in most cases. This indicates that the models can learn generic knowledge from MIMIC and help with predictions on the HDRJH dataset. These models were further ensembled by taking the average, and the ROC curves are shown in Figure S4.

The effect of transfer learning on HDRJH dataset sampled at different ratios are shown in Figure S5-8.

Table S4. **The results of the ensemble model on the HDRJH dataset**

| **Preceding hours** | Accuracy | AUC | Sensitivity | Specificity |
| --- | --- | --- | --- | --- |
| 1 | 0.86 | 0.94 | 0.72 | 1 |
| 2 | 0.86 | 0.94 | 0.72 | 1 |
| 3 | 0.88 | 0.94 | 0.75 | 1 |
| 4 | 0.82 | 0.94 | 0.64 | 0.99 |
| 5 | 0.87 | 0.94 | 0.75 | 0.99 |

Table S5. **The results of LightGBM on the HDRJH dataset**

| **Preceding hours** | Transfer learning | Training set | Accuracy | AUC |
| --- | --- | --- | --- | --- |
| 1 | N | MIMIC | 0.74 | 0.85 |
| 2 | N | MIMIC | 0.73 | 0.83 |
| 3 | N | MIMIC | 0.72 | 0.84 |
| 4 | N | MIMIC | 0.74 | 0.86 |
| 5 | N | MIMIC | 0.69 | 0.82 |
| 1 | N | HDRJH | 0.8 | 0.94 |
| 2 | N | HDRJH | 0.81 | 0.92 |
| 3 | N | HDRJH | 0.81 | 0.92 |
| 4 | N | HDRJH | 0.81 | 0.92 |
| 5 | N | HDRJH | 0.81 | 0.94 |
| 1 | Y | MIMIC+HDRJH | 0.82 | 0.93 |
| 2 | Y | MIMIC+HDRJH | 0.83 | 0.93 |
| 3 | Y | MIMIC+HDRJH | 0.85 | 0.93 |
| 4 | Y | MIMIC+HDRJH | 0.83 | 0.93 |
| 5 | Y | MIMIC+HDRJH | 0.87 | 0.94 |

Table S6. **The results of MLP on the HDRJH dataset**

| **Preceding hours** | Transfer learning | Training set | Accuracy | AUC |
| --- | --- | --- | --- | --- |
| 1 | N | MIMIC | 0.76 | 0.87 |
| 2 | N | MIMIC | 0.72 | 0.84 |
| 3 | N | MIMIC | 0.75 | 0.85 |
| 4 | N | MIMIC | 0.76 | 0.86 |
| 5 | N | MIMIC | 0.75 | 0.86 |
| 1 | N | HDRJH | 0.84 | 0.92 |
| 2 | N | HDRJH | 0.84 | 0.92 |
| 3 | N | HDRJH | 0.83 | 0.91 |
| 4 | N | HDRJH | 0.84 | 0.92 |
| 5 | N | HDRJH | 0.83 | 0.92 |
| 1 | Y | MIMIC+HDRJH | 0.87 | 0.93 |
| 2 | Y | MIMIC+HDRJH | 0.88 | 0.93 |
| 3 | Y | MIMIC+HDRJH | 0.86 | 0.92 |
| 4 | Y | MIMIC+HDRJH | 0.78 | 0.93 |
| 5 | Y | MIMIC+HDRJH | 0.85 | 0.93 |

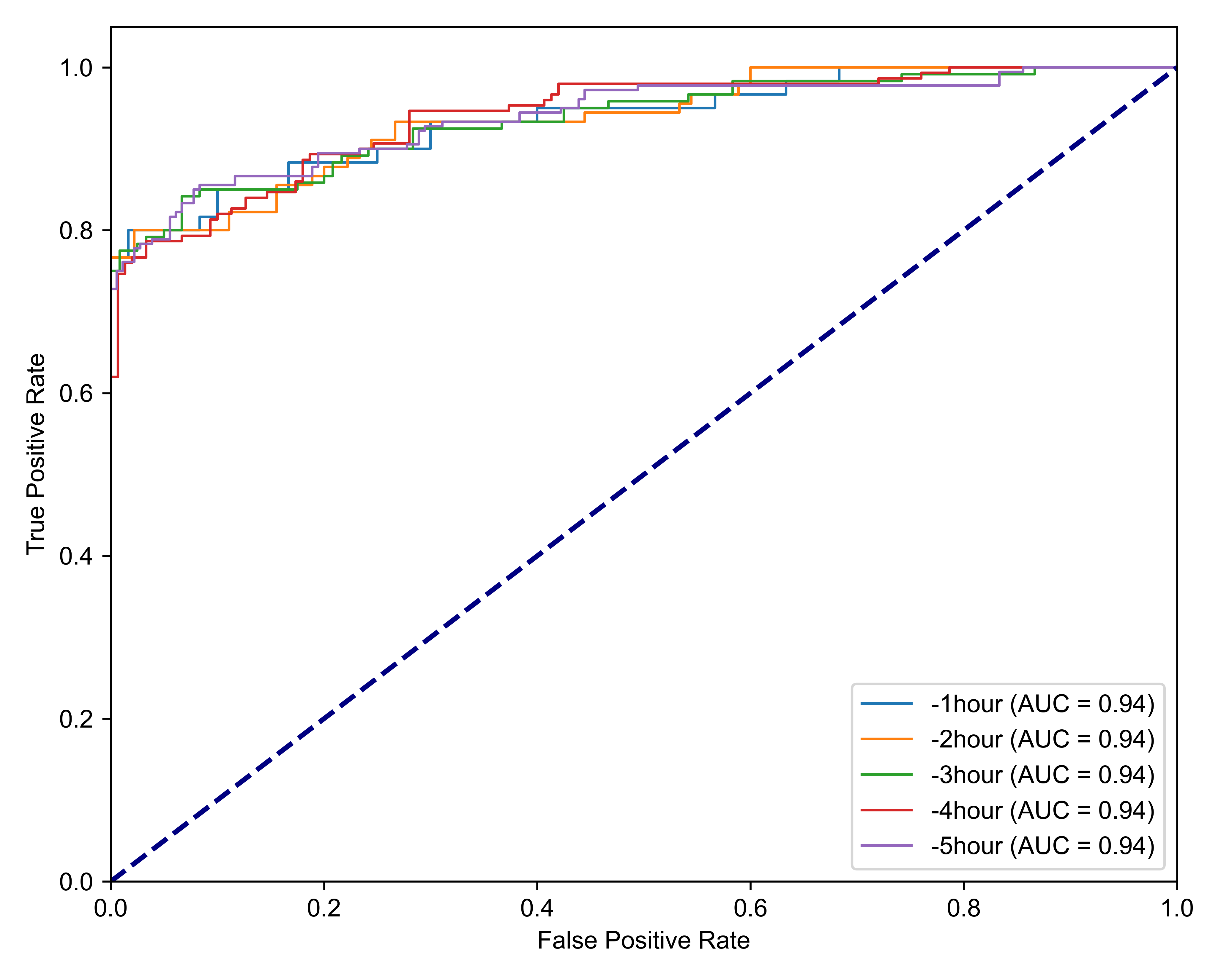

Figure S4. **ROC curves of ensemble models detecting sepsis in 1–5-h preceding on the HDRJH dataset.**

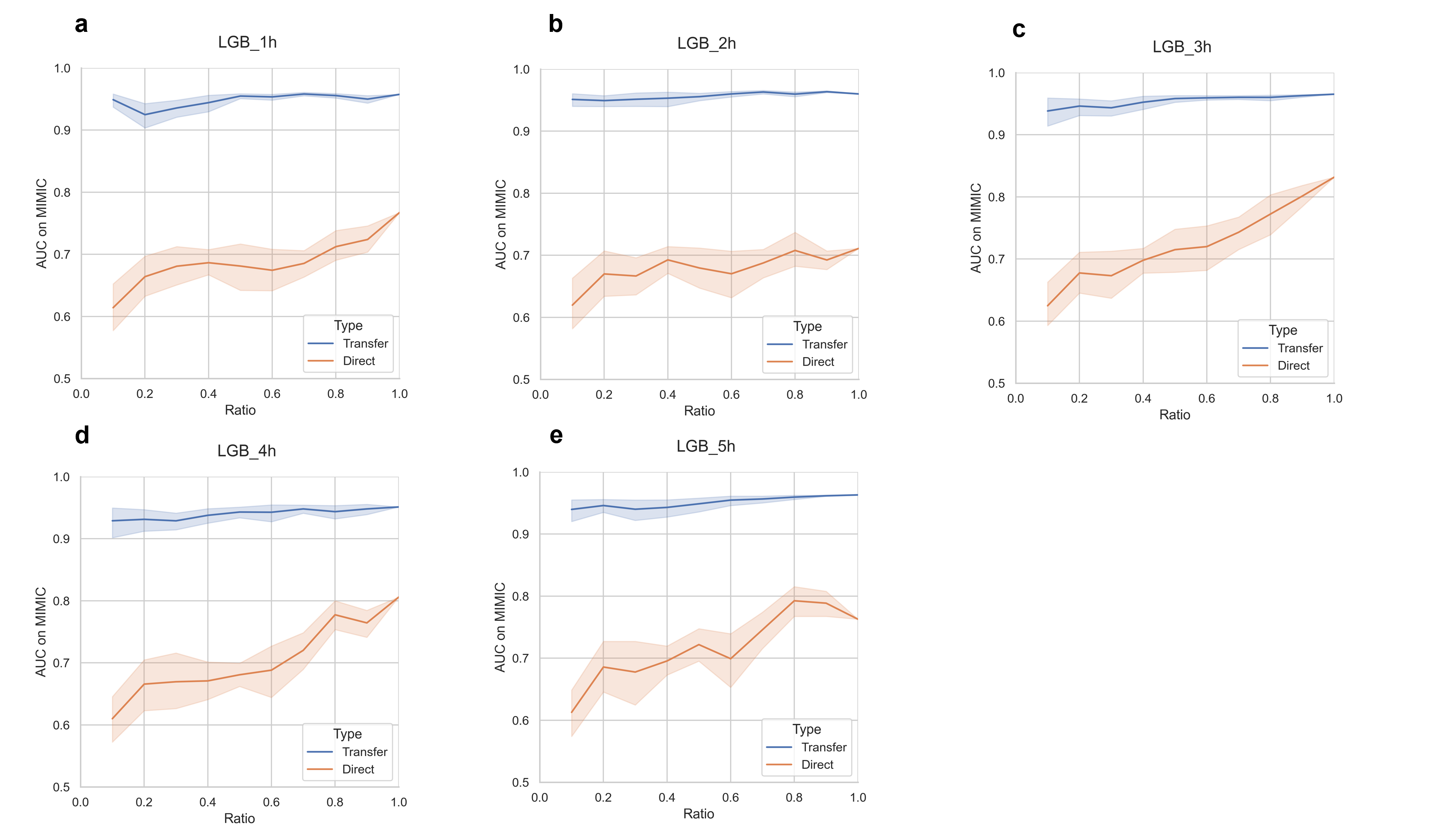

Figure S5. **The AUCs of LightGBM models trained on sampled HDRJH dataset and tested on MIMIC.**

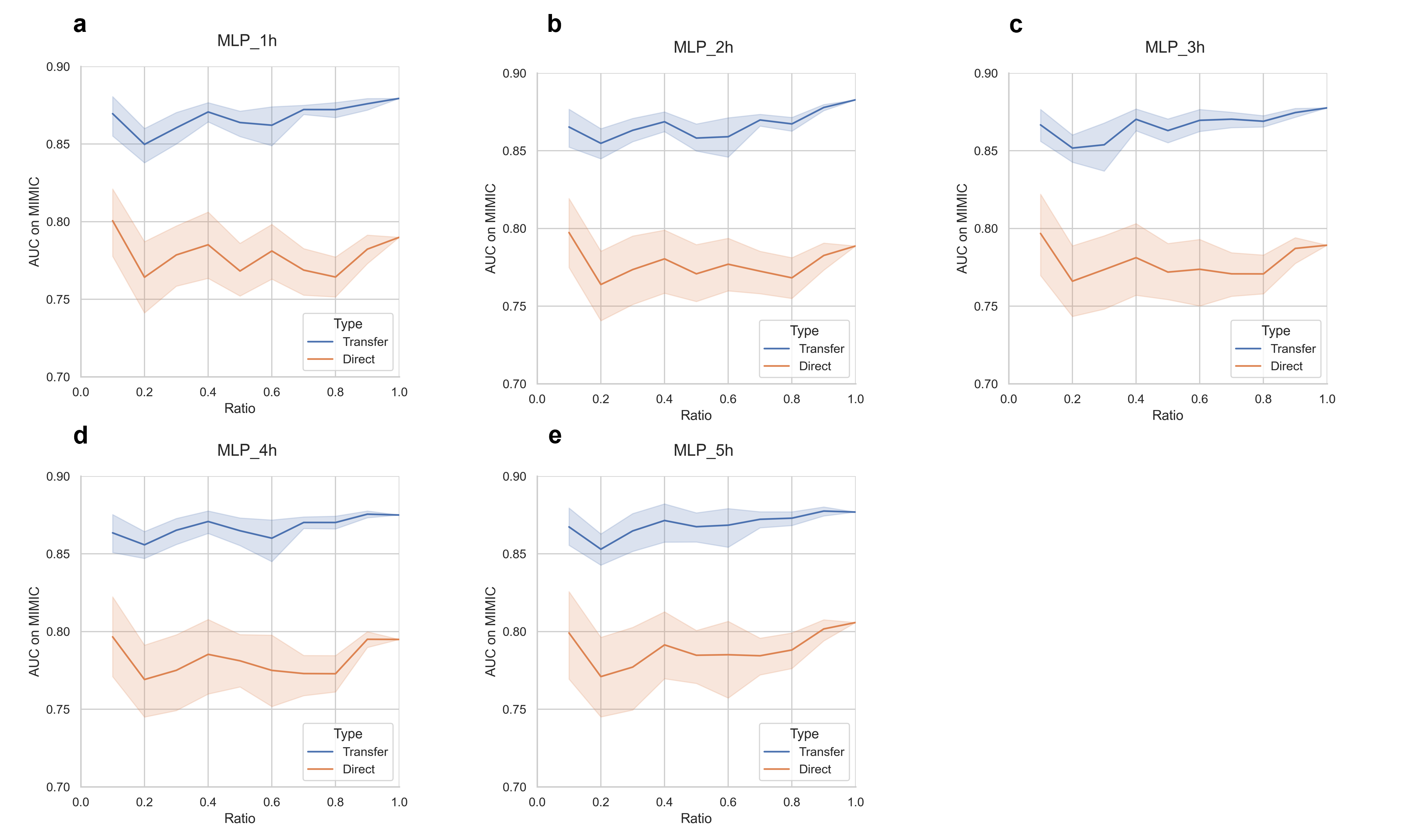

Figure S6. **The AUCs of MLP models trained on sampled HDRJH dataset and tested on MIMIC.**

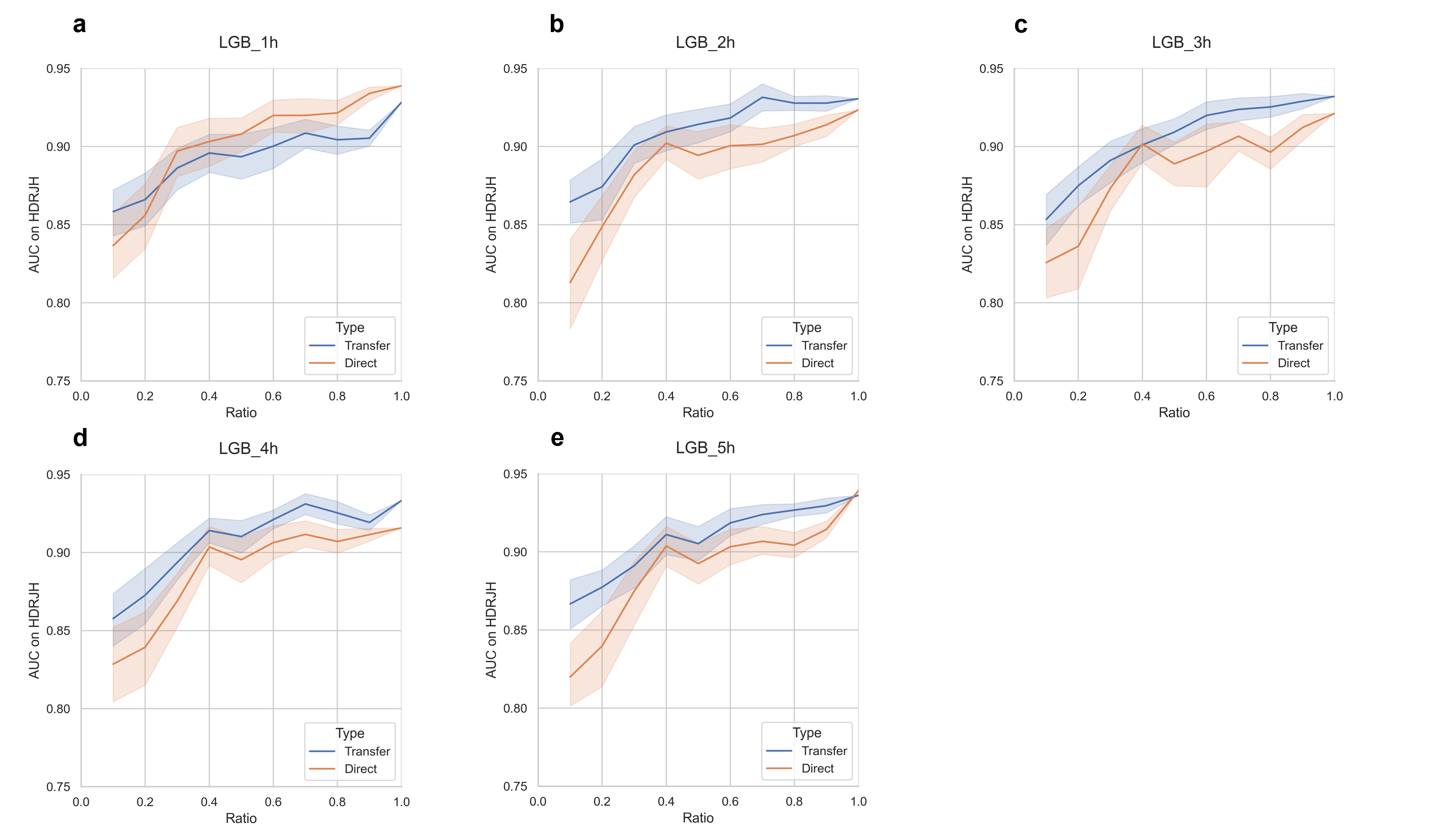

Figure S7. **The AUCs of LightGBM models trained on sampled HDRJH dataset and tested on HDRJH.**

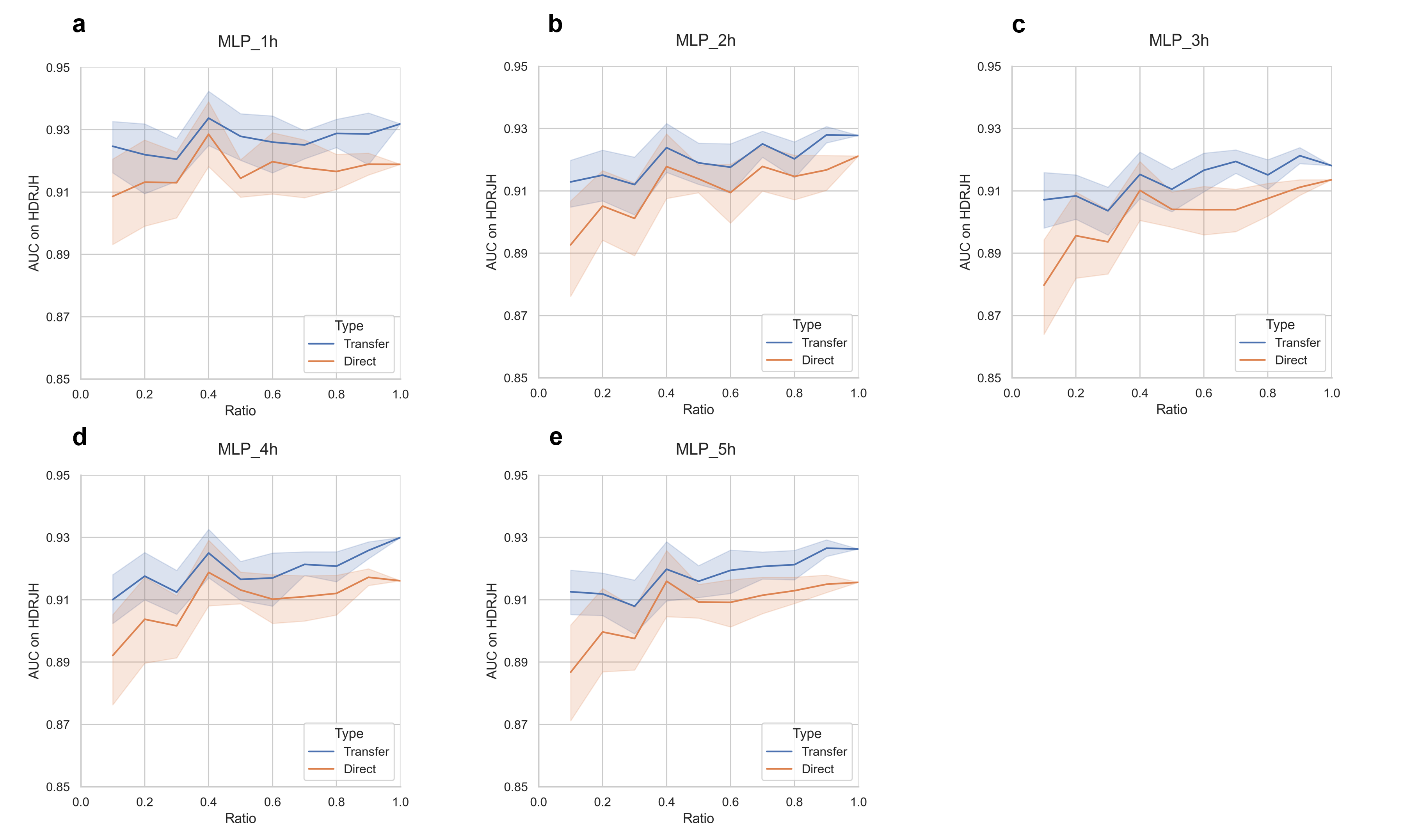

Figure S8. **The AUCs of MLP models trained on sampled HDRJH dataset and tested on HDRJH.**

### Appendix 9 Feature importance

We used the Shapley Additive Explanations (SHAP) analysis to explore the importance of features of different models. The SHAP value of each feature represents the impact of the feature on the output of the model. For LightGBM models trained on the MIMIC-III dataset, the beeswarm plots were shown in Figure S9. The importance of features of the same hour and from the same variable were combined, and the twenty most important features were displayed in the figures. As shown in Figure S9, several features were found to be important in most models, e.g., FiO_2_, fibrinogen, calcium, ventilation, age, albumin, antibiotics, O_2_Sat, PCO_2_, peak inspiratory pressure, and SOFA.

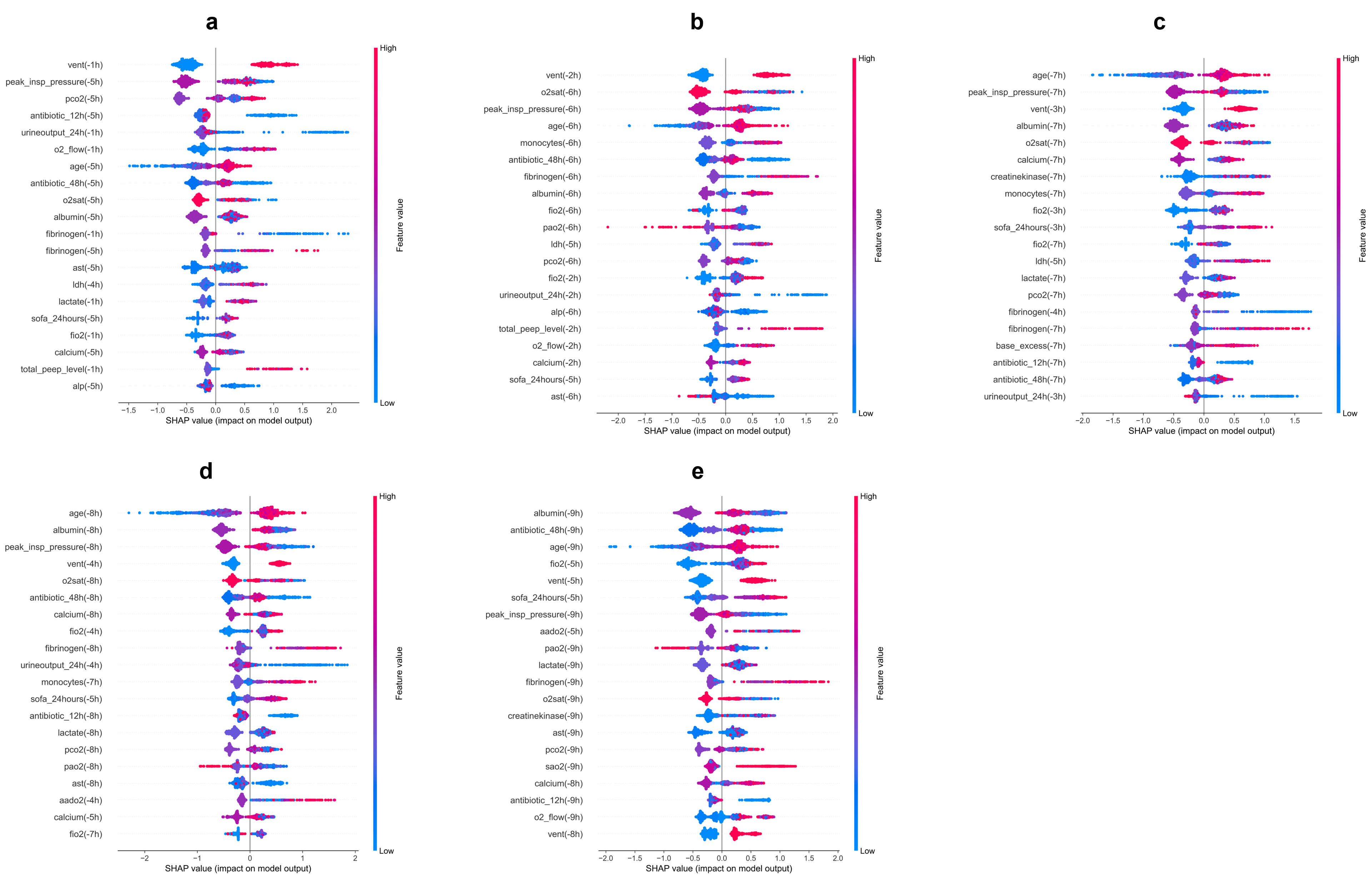

Figure S9. **Feature importance of LightGBM models on the MIMIC-III dataset.** a - e are the beeswarm plots of the models detecting sepsis in 1 - 5 hours preceding, respectively. In each subgraph, Y-axis represents different features and X-axis represents the SHAP value of the sample point represented by one dot. A vertical thickness of a dot cluster represents the number of dots that fall on this SHAP value. The color of a dot represents the value of the feature for one sample, with blue indicating low and red indicating high. The names of features on the Y-axis follow the format ‘feature (hours before onset)’.

For the models in SEPRES, the feature importance for LightGBM and MLP models was calculated separately in Figure S10 and Figure S11. The feature importance of LightGBM and MLP models was not consistent overall, although they both yielded high AUCs. For LightGBM models, antibiotics, respiratory rate, total PEEP level, fibrinogen, temperature, net balance, and age were found to be important in most models. For MLP models, age, respiratory rate, ventilation, heart rate, antibiotics, and temperature were found to be important in most models.

Some of these features (antibiotics, respiratory rate, temperature, ventilation, heart rate) were related to the definition of Sepsis-3 or SIRS, while there was also literature arguing for an association between some of these features (respiratory rate [5], fibrinogen [6], net balance [7], and age [8]) and the severity or the mortality of sepsis.

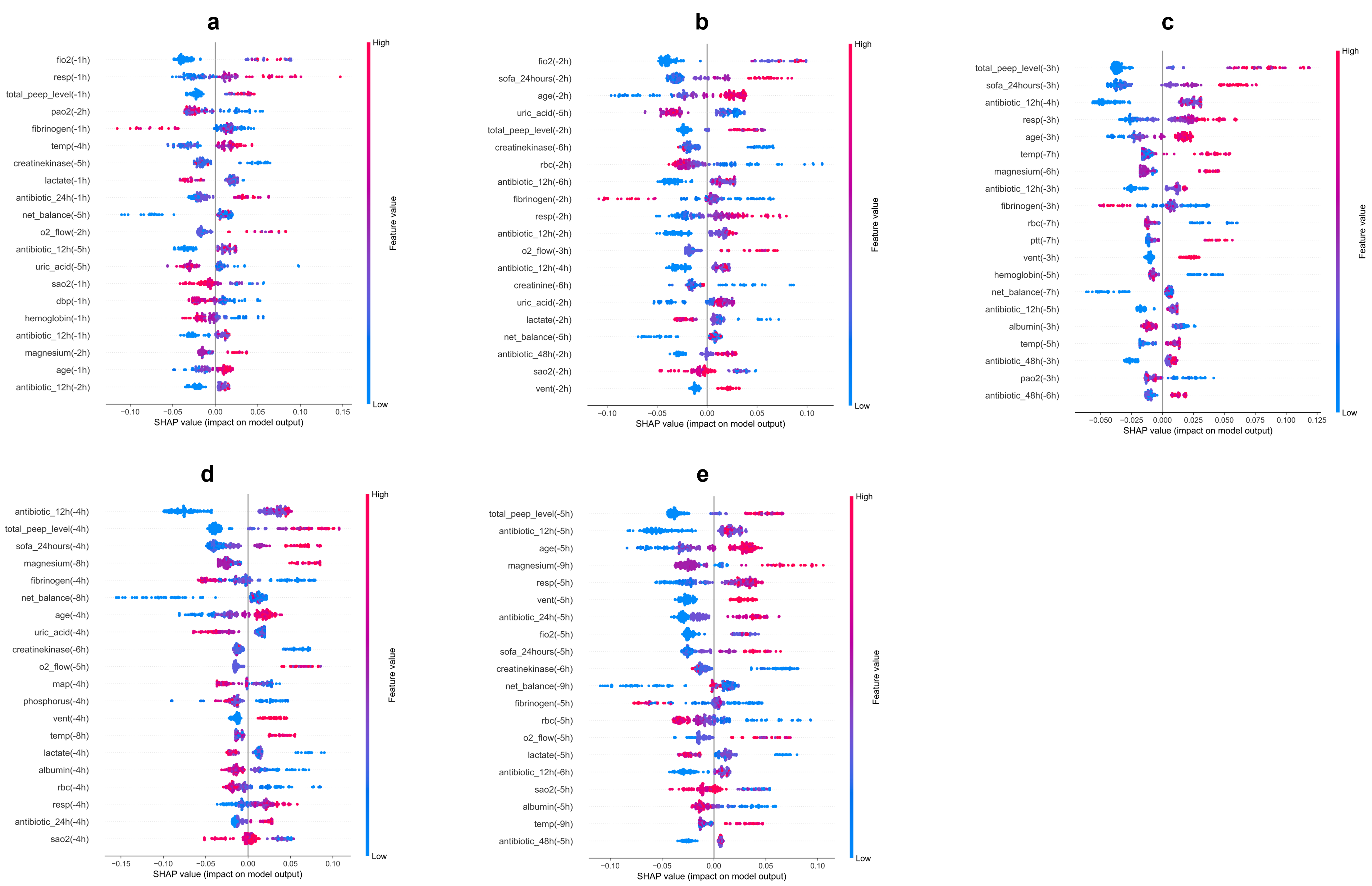

Figure S10. **Feature importance of LightGBM models in the sepsis early warning module.** a - e are the beeswarm plots of the models detecting sepsis in 1 - 5 hours preceding, respectively.

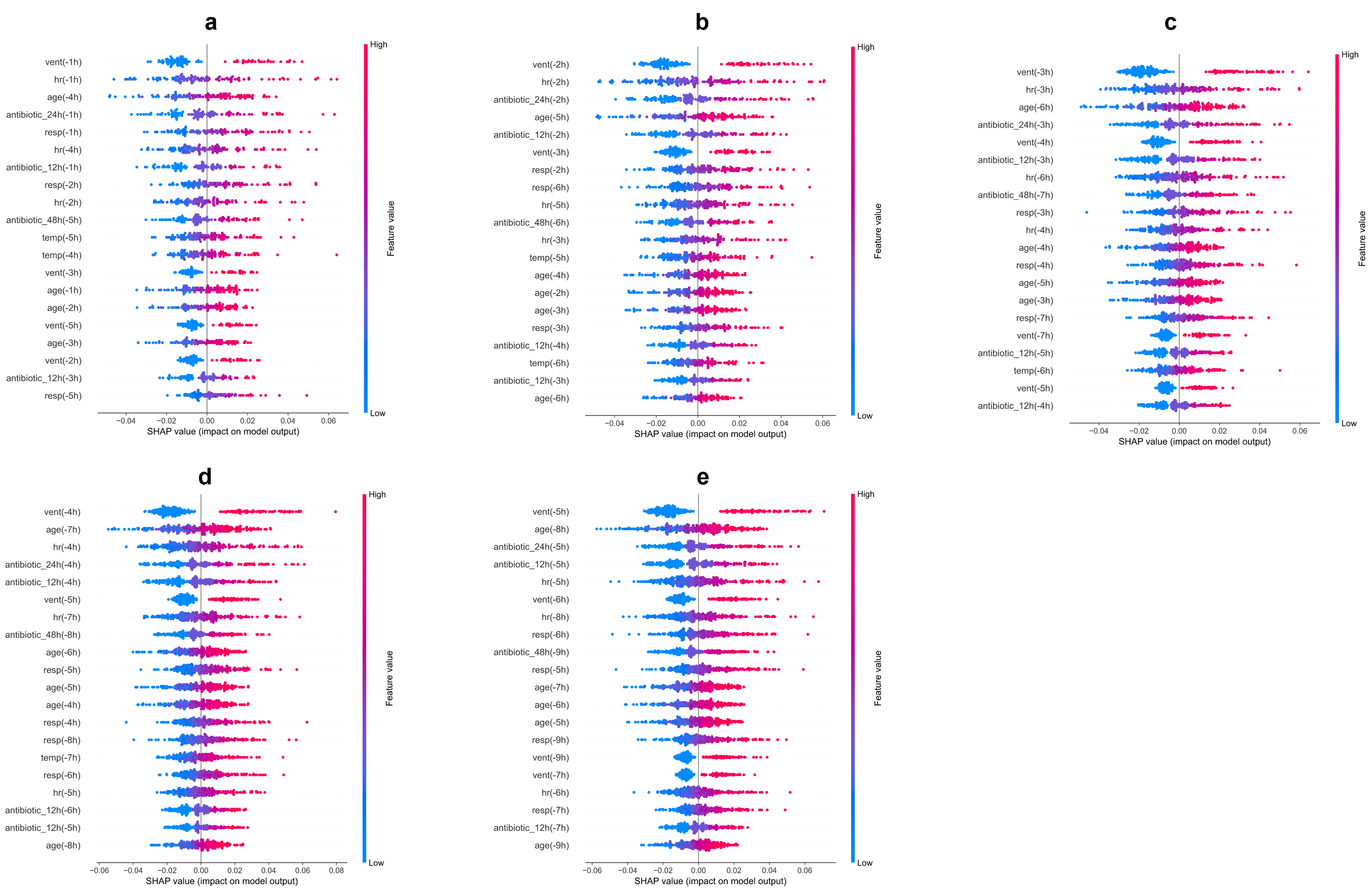

Figure S11. **Feature importance of MLP models in the sepsis early warning module.** a - e are the beeswarm plots of the models detecting sepsis in 1 - 5 hours preceding, respectively.

### Appendix 10 Case studies

In the actual operation at Ruijin Hospital, the threshold was increased from the default 0.5 to 0.7 in order to reduce the false alarm rate of sepsis warning. The adjusted specificity was increased to about 0.88, although the sensitivity was reduced to about 0.72, which is shown in Table S7.

Table S7. **The results of real-time data with threshold at 0**.**7**

| **Preceding hours** | Accuracy | AUC | Sensitivity | Specificity |
| --- | --- | --- | --- | --- |
| 1 | 0.75 | 0.86 | 0.69 | 0.89 |
| 2 | 0.78 | 0.88 | 0.74 | 0.89 |
| 3 | 0.77 | 0.9 | 0.73 | 0.89 |
| 4 | 0.77 | 0.9 | 0.73 | 0.89 |
| 5 | 0.79 | 0.89 | 0.75 | 0.89 |

**Positive case:**

A man aged 41-50 was transferred to our ICU due to intestinal obstruction and abdominal infection after the pancreaticoduodenectomy (PD) one month ago. After the drainage surgery, the condition of the patient aggravated in the early morning of February 15, 2021, with multiple organ dysfunction. At 12:00 AM, the SOFA score was 13 (ΔSOFA ≥ 2 within 72 hours). In combination with the suspected infection, the patient was diagnosed as sepsis according to the definition of Sepsis-3. Our sepsis early prediction model has already predicted the incidence of sepsis three hours in advance. At 9:00 AM, the sepsis early prediction model showed that the confidence index (CI) of sepsis incidence in one hour was 0.72 which was over the prediction threshold 0.7 (Figure S12), and the prediction software at the medical terminal UI interface raised the pre-warning prompt. The high CI at 10:00 AM and 11:00 AM indicated the aggravation of the patient. The early prediction of sepsis occurrence by our model effectively guided medical practitioners to appropriately pay more attention to this patient, thereby leading to the early diagnosis of sepsis.

**Negative case:**

A man aged 41-50 was admitted to our hospital due to chronic renal failure (uraemia period with acute exacerbation) in combination with metabolic acidosis and renal failure after renal transplantation as well as severe hypertension. The patient was given RRT hypertension control treatment. During the monitoring in the ICU, the SOFA score was high (7.0), and the condition of the patient was severe due to several comorbidities and complications in combination with uraemia. However, there was no evidence of ΔSOFA ≥ 2 within 72 hours. Consistently, the CI of sepsis incidence were all lower than the warning threshold 0.7, suggesting no sepsis occurrence after admission. Therefore, this is a negative case.

**False positive case:**

A man aged 81-90 was admitted into our hospital due to intracranial space occupying lesion (frontotemporal malignant tumor). The patient was transferred to the ICU due to hospital-acquired pneumonia (klebsiella pneumoniae) and respiratory failure after surgery. The condition of the patient was severe with fluctuations in body temperature during the monitoring period. The SOFA score was stable at 6.0 without the evidence of ΔSOFA ≥ 2 within 72 hours, suggesting no sepsis occurrence. However, the CI of sepsis incidence in 5 hours predicted by our sepsis early prediction model was over the warning threshold 0.7. Therefore, this is a false positive case.

The negative control group used during training the sepsis early prediction model was non-septic patients. Therefore, the prediction model may falsely determine the patient that is severely ill to be sepsis. This is a limitation of our present model. To solve this problem, we will analyze the control group by delaminating patients with different severities and further optimize the prediction model.

**False negative case:**

A man aged 61-70 was admitted into our hospital due to sellar tumor. During the transnasal transsphenoidal resection of pituitary adenoma, the patient had hemorrhagic shock and hypoxic-ischemic encephalopathy. The patient was transferred into the ICU and was given ventilation and anti-infectious therapy. The SOFA score showed an increase from 6 to 9 (ΔSOFA ≥ 2 within 72 hours) at 06:00 PM. In combination with the evidence of infection, the patient was diagnosed as sepsis. However, as shown on the medical terminal interface, the CI of sepsis incidence was below the warning threshold 0.7. Therefore, this is a false negative case.

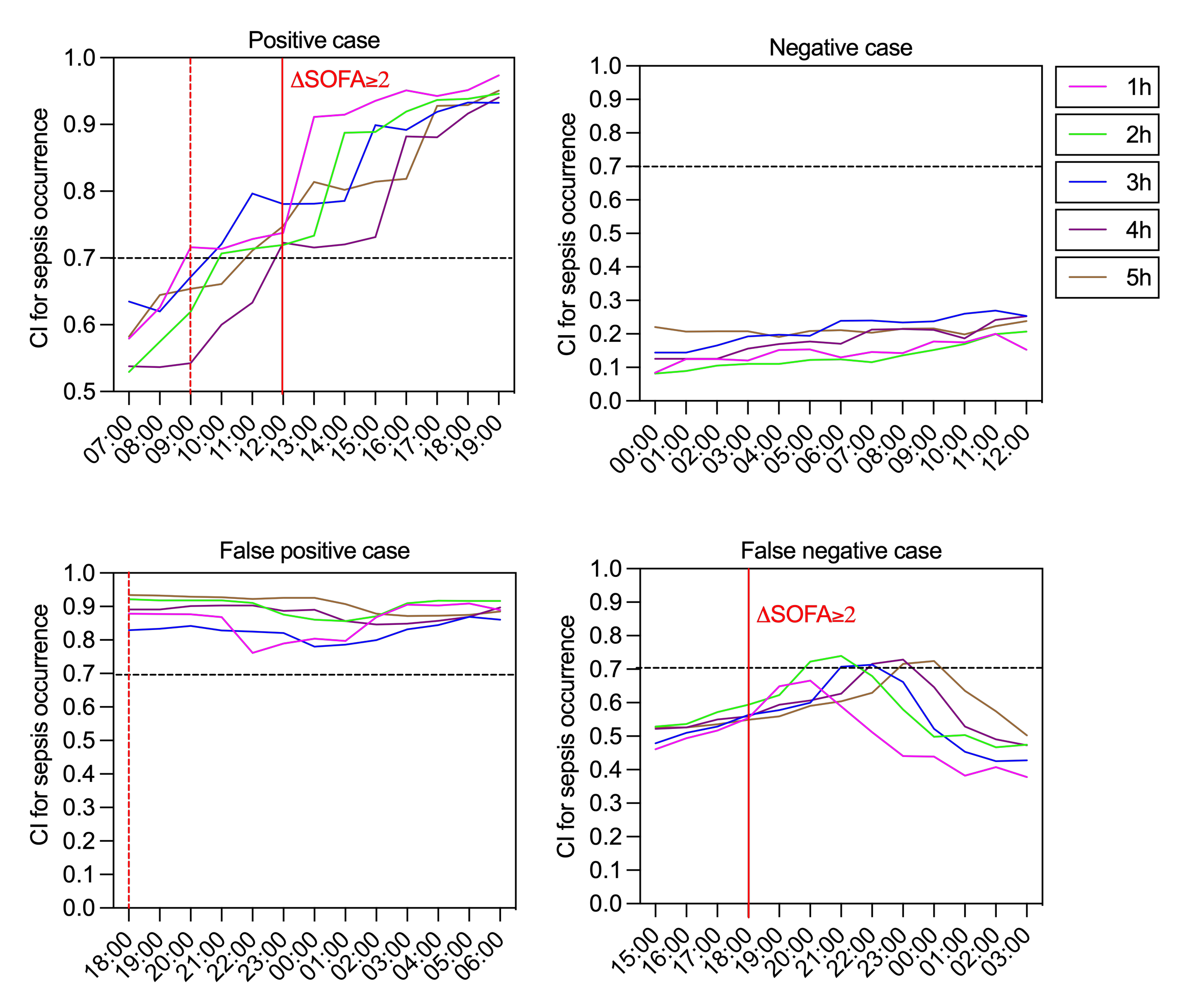

Figure S12. **The confidence index (CI) of sepsis prediction of the patient at each time node.**

We concluded that the ICU duration for all these three false negative cases was relatively short, therefore the data collected for prediction was limited, which may lead to inaccurate prediction for sepsis occurrence. As supporting evidence, there were only 17 positive cases with an onset within 2 days in the HDRJH dataset, which may not be sufficient for the model to learn positive cases in the absence of enough data.

As a further experiment, we selected cases in the HDRJH that were screened out due to too few valid variables, and the performance on these cases is shown in Table S8. It can be seen that the sensitivity produced a substantial decrease in sensitivity in contrast to a slight decrease in specificity. This suggests that when there is insufficient valid data for a case, the model will tend to produce lower CI of sepsis incidence, resulting in false negative cases. This may need to be addressed by the inclusion of more positive cases with fewer valid variables, as well as data augmentation.

Table S8. **The results on cases with few valid variables**

| **Preceding hours** | Accuracy | AUC | Sensitivity | Specificity |
| --- | --- | --- | --- | --- |
| 1 | 0.63 | 0.65 | 0.32 | 0.95 |
| 2 | 0.63 | 0.67 | 0.32 | 0.94 |
| 3 | 0.63 | 0.67 | 0.33 | 0.94 |
| 4 | 0.64 | 0.67 | 0.32 | 0.95 |
| 5 | 0.63 | 0.69 | 0.31 | 0.95 |

### Reference

1 Nemati S, Holder A, Razmi F, Stanley MD, Clifford GD, Buchman TG (2018) An interpretable machine learning model for accurate prediction of sepsis in the ICU. Crit Care Med 46:547. https://doi.org/10.1097/CCM.0000000000002936
2 Moor M, Horn M, Rieck B, Roqueiro D, Borgwardt K (2019) Early recognition of sepsis with Gaussian process temporal convolutional networks and dynamic time warping. Machine Learning for Healthcare Conference 2−26. PMLR.
3 Hong LK, Wogan G, Vacca L, Tidor B, inventors. Peach IntelliHealth Pte Ltd., assignee (2019) System and method for predicting sequential organ failure assessment (sofa) scores using artificial intelligence and machine learning. United States Patent 20190259499.
4 Silva Á, Cortez P, Santos MF, Gomes L, Neves J (2008) Rating organ failure via adverse events using data mining in the intensive care unit. Artif Intell Med 43:179-93. https://doi.org/10.1016/j.artmed.2008.03.010
5 Kenzaka T, Okayama M, Kuroki S, et al (2012) Importance of vital signs to the early diagnosis and severity of sepsis: Association between vital signs and sequential organ failure assessment score in patients with sepsis. Intern Med 51:871−876. https://doi.org/10.2169/internalmedicine.51.6951
6 Matsubara T, Yamakawa K, Umemura Y, et al (2019) Significance of plasma fibrinogen level and antithrombin activity in sepsis: A multicenter cohort study using a cubic spline model. Thromb Res 181:17−23. https://doi.org/10.1016/j.thromres.2019.07.002
7 Brotfain E, Koyfman L, Toledano R, et al (2016) Positive fluid balance as a major predictor of clinical outcome of patients with sepsis/septic shock after ICU discharge. Am J Emerg Med 34:2122−2126. https://doi.org/10.1016/j.ajem.2016.07.058
8 Martin GS, Mannino DM, Moss M (2006) The effect of age on the development and outcome of adult sepsis. Crit Care Med 34:15−21. https://doi.org/10.1097/01.CCM.0000194535.82812.BA

1. ^*^: adjusted p-value (between MIMIC and HDRJH) < .05

   ^**^: adjusted p-value (between MIMIC and HDRJH) < .01

   ^***^: adjusted p-value (between MIMIC and HDRJH) < .001
